# Delayed Discharges: Does Staff Well-Being Matter?

**DOI:** 10.1101/2020.06.10.20127522

**Authors:** Manhal Ali, Reza Salehnejad

## Abstract

Delayed discharges of patients from hospitals, also known as “bed–blocking” is a long standing policy concern. Such delays can increase hospital treatment costs and may also lead to poorer patient health and experience. Prior research indicates that external factors, such as, greater availability and better affordability of long term care associated with lower delays. Using theories from Economics, this study examines the role of within–hospital factors, namely, staff well–being in alleviating hospital delayed days. We use a new panel database of delays in all English hospital trusts from 2011/12 to 2014/15. Employing longitudinal count data models, the paper finds that staff well–being is associated with lower hospital delayed discharges controlling for long–term factors and management quality. The findings are robust to alternative methods and measures of delayed discharges.

## 1 Introduction

Delayed discharges or transfers^1^ of care also termed as “bed–blocking” occurs when an adult inpatient in a hospital is medically optimised to go home or move to a less acute stage of care, but is prevented from doing so. Since 2010/11, there has been a rising upward trend in the number of delays in patients being discharged from hospital (figure 1).

**Figure 1:**
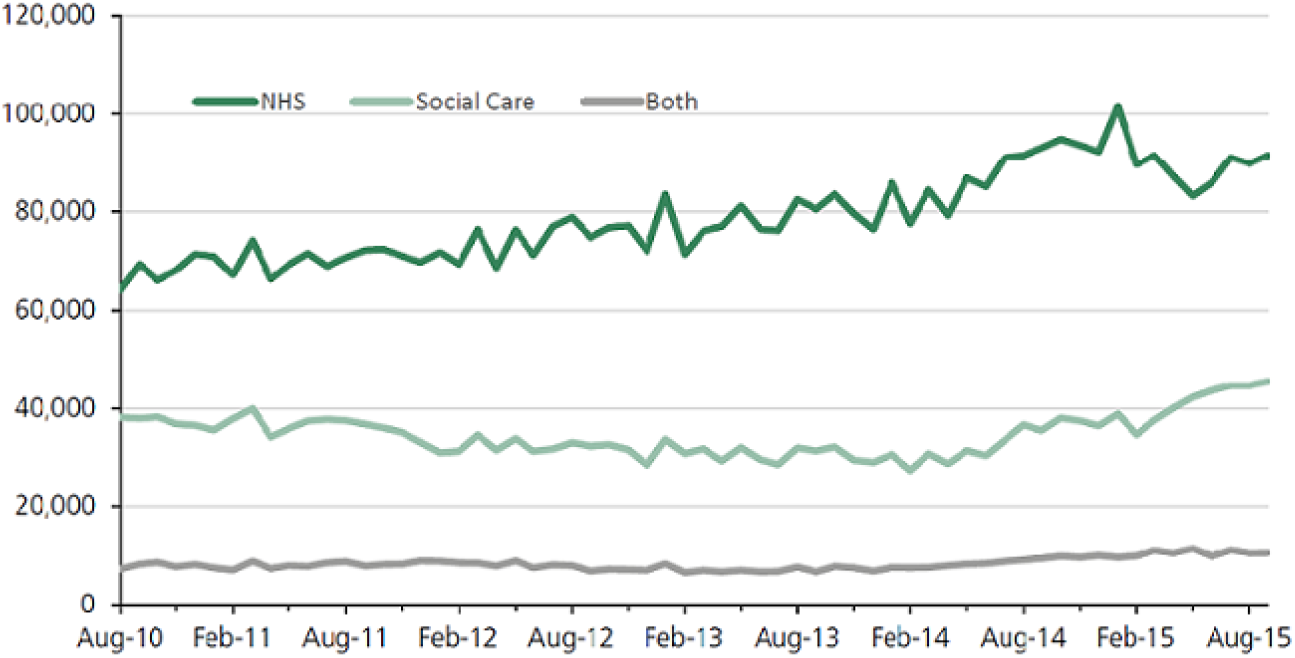
Delayed Days over 2010–2015 (Bates 2015)

Between 2011/12 and 2014/15, the number of bed days used by patients who were delayed grew by 60 per cent (Edwards, 2017). There were 1.2 and 1.6 million bed–days lost in 2013/14 and 2014/15 respectively with an average of around 4,500 per day in 2014/15 (Gaughan et al., 2016). Such delays not only pose significant financial challenges to hospitals, but also have adverse clinical consequences for the patients (Bryan et al., 2006; Gaughan et al., 2015; Hendy et al., 2012; Lewis and Purdie, 1988). Hospital delayed discharges for patients remains a significant policy concern not only for the NHS but also for health–care sectors of other OECD countries. In NHS England, delays can either be attributed to social care or local authority in which the patient resides or to NHS i.e. hospitals.^2^ Majority of the delays are however due to NHS where waiting for further intermediate care and medical tests remain the major causes for delays.

Since hospital care is relatively more expensive than alternative forms of care such as nursing or residential home care, delayed discharges is a signal of allocative inefficiency. It also has productivity implications. Firstly, delayed transfers of patients to their residence or alternative forms of care can lead to loss of hospital bed–days, thereby reducing the efficiency on how inputs are used. Secondly, “bed–blocking” may stymie the number of patients seen, investigated or treated and therefore affecting hospitals’ revenue and output activity. Hence, delayed discharges can serve as a measure of productivity—reduction in delayed days are interpreted as increasing productivity.

Prior studies have attempted to explain rising delays in discharging patients and variations across healthcare providers by using internal, hospital–organisational and external factors such as availability of alternative or long–term care services. A lot of emphasis is however placed on social care expenditure and the availability of long term care services for the recent problems of rising and persistent patient delayed days (Gaughan et al., 2017, 2015, 2016; Manzano-Santaella, 2010). This paper focuses on the role of hospitals internal factors in the form of staff well–being and analyse its relationship with hospital delayed discharges and, examines the extent of its efficacy in reducing discharge rates.^3^

This paper argues that in addition to the external factors, problems of delayed transfers is equally if not more related to hospitals own internal processes or systems where human capital is at the forefront. These internal system inefficiencies amongst others comprises of failures in hospitals’ discharge planning process, medical errors, lack of (skilled) staff or rehabilitation services, waiting for another opinion, a planned investigation or decision from another consultant. Since staff are so integral for innovation and sustainability of an organisation according to human capital and other related theories, this paper builds and tests whether staff–well being is beneficial to hospitals value in helping to reduce delayed transfers of care for NHS patients. There is a fairly large literature that studies hospital delayed transfers of care. One group of literature study the causes of delays at a single particular hospital or unit, for instance critical care, surgical or rehabilitation unit and/or using detailed patient–level and organisational data (Hendy et al., 2012; Lin et al., 2009; Majeed et al., 2012; Wong et al., 2009). Whereas, a second group of literature study delayed days across a handful of healthcare providers or hospitals using interviews (Baumann et al., 2007) or detailed clinical data (Challis et al., 2014). Other form of literature, for example Godden et al. (2009), examines trends in delayed activity from 2001/02 to 2006/07 using routine hospital data and, evaluated the impact of Community Care Act 2003 in raising efficiency across health and social care. Godfrey and Townsend (2009) compares and examines the different policy approaches implemented in England and Scotland in reducing delayed discharges. Similarly, McCoy et al. (2007) analysed trends in delayed discharges from 2003/04 to 2005/06 using routine and survey data from 150 social service departments in England.

Comparatively, there exists a very small amount of studies in the health economics literature that studied hospital delayed discharge rates exclusively. In one of the few studies using NHS Local Authority (LA) level data, Gaughan et al. (2015) investigates the extent to which nursing home beds and prices reduce bed–blocking. Their findings suggest that delayed discharges respond to availability of care home beds but with modest effects. In addition, they also find evidence of spill–over effects across LAs: more care home beds or fewer patients aged over 65 years in nearby LAs are associated with fewer delayed discharges. Using two–year data from 1998/99 to 1999/00 on 150 LAs, Fernandez and Forder (2008) find that English patients living in LAs with fewer care home and nursing beds were more likely to have a delayed discharge. Similar findings are also reported in Forder (2009). A recent study by Gaughan et al. (2016) are amongst the first to examine variations in delayed discharges across hospitals. Using a 3 year panel data from 2011/12 to 2013/14 and employing various case–mix, long term, quality and hospital characteristics, they find that hospital’ s with Foundation Trust status are less likely to experience delayed days compared to non–Foundation Trust hospitals. Moreover, they also find hospitals’ in regions with high availability of long term care beds and hospital’ s with mental health statuses are also less likely to experience delayed days of its patients.

According to happy–productive worker thesis (Brief, 1998; Spector, 1997), the tendency of happy people to emphasize the positive aspect of their job leads to higher job performance like enhanced productivity. This in turn improves overall organisational performance. To this end, the economics literature has been accumulating evidence by examining the link between staff satisfaction and firm outcomes, for instance in finance (Edmans, 2011, 2012; Edmans et al., 2014; Huang et al., 2015) and in manufacturing (Boö ckerman and Ilmakunnas, 2012). Using instrumental variable techniques and data from Finnish manufacturing plants from the period 1996–2001, Boö ckerman and Ilmakunnas (2012) finds a compelling evidence on the link between employee satisfaction measured on a six–point scale and establishment–level productivity. Similar evidences are also found in Bryson et al. (2015), Kruse et al. (2012), Best (2008) and Ahluwalia (2015).

This thesis especially holds in jobs at an organisation such as hospital that requires continuous interactions with co–workers and patients. There also exists studies from the healthcare literature that finds evidence of association between various measures of staff satisfaction and hospital performances measured in terms of quality such as mortality (Bhatnagar et al., 2012; Chang et al., 2017; Lowe, 2012; Peltier et al., 2009; Pinder et al., 2013; Powell et al., 2014), finances (Harmon et al., 2003; Peltier et al., 2009; West et al., 2011), waiting times (Chang et al., 2017), absenteeism and turnover (Bhatnagar et al., 2012; Chang et al., 2017; Powell et al., 2014). For instance, Pinder et al. (2013) finds that higher hospital staff satisfaction for medical and nursing staff are associated with lower hospital standardised mortality. Most of these studies however, use cross–sectional based designs and lack statistical sophistication that are necessary to find a more robust relationship. Results may also by biased if the issue of reverse causality or simultaneity is not taken into account. Results will be biased if hospital performances such as mortality or patient experiences affects staff morale and satisfaction. Results may also be biased if the exogeneity assumption is violated.

This paper aims to test the happy–productive worker hypothesis with respect to hospital delayed transfers of care. If the theoretical predictions of happy–productive worker and related theories hold, we expect that greater welfare of hospitals’ staff will help to alleviate patients’ delayed transfer of care from the hospitals via enhanced effort and productivity. To allow for the possibility of misclassification, the paper has estimated models for days delayed that are due to NHS (i.e. hospitals) and both NHS and social care. In addition, this paper also looks at disaggregated causes of delays and estimated models of delayed transfers due to major causes. The measure of staff–satisfaction for NHS hospitals are added for the following staff categories: clinical/medical, nursing and total (i.e. all clinical and non–clinical staff combined). There is a growing and a strong body of studies including the earlier chapters that has found that management matters in healthcare. To prevent potential confounding, this paper therefore examines the relationship between delayed days and well–being by including management quality at each hospital as an additional covariate.

By using a panel sample between 2011/12–2014/15, this paper finds that there is a significant and positive association between staff satisfaction and lower delayed days even after controlling for hospital and year fixed effects and other covariates. The results overwhelmingly find consistent and significant support for the well–being of hospitals’ clinical staff in alleviating hospital delayed discharge rates. The results are robust to various alternative specifications and measures and, the paper took steps to address some of the econometric issues such as endogeneity.

The results are consistent with the human capital theories which argue that employee satisfaction causes stronger firm performance through improved recruitment, retention, and motivation. The results are also consistent with some of the other recent theories of the firm that focused on employees as the key assets such as Rajan and Zingales (1998), Carlin and Gervais (2009) and Berk et al. (2010).

The findings have implications for both NHS managers and policy makers. Exploratory analysis suggests that there exists significant and persistent variations in employee satisfaction scores amongst the clinical staff of NHS hospitals. Therefore, potential may exist where managers could incorporate HR practices and policies or create an environment that seek to improve staff morale and welfare.

Using relevant theories, a conceptual framework is provided that links staff well–being to higher hospital performances in terms of lower days delayed. It is believed that this is the first study in the literature that attempts to examine variations in delayed days across hospitals by focusing on its internal factors in the form of staff well–being.

The rest of the paper is organised as follows. Section 2 provides the theoretical framework and the main hypotheses. Section 3 explains the relevant data and provide summary statistics. Section 4 introduces the analytical approach of this paper. And section 5 presents the main empirical results and detailed robustness tests. The last two sections discusses the findings of this paper, policy implications, limitations and present the conclusion.

## 2 Theoretical Framework

### 2.1 The Conceptualisation of Well–Being

The standard neo–classical theory of labour supply assume that individuals’ utility is a function of income and leisure. Income is generated from work, but this eats into time that is available for leisure. Therefore, individuals’ make trade–off decisions between income and leisure in order to maximise their utility. In this view, when holding income constant, additional time devoted to work create disutility. It follows that when an individual becomes unemployed, the pain inflicted by loss of income should be adjusted due to gain in leisure time. Research on well–being appears to contradict this, however. It indicates that, holding income constant, work makes a positive contribution to overall life satisfaction and general happiness that is substantial and positive (Bryson and MacKerron, 2017).

Paid work is an important part of many people’ s life. They spend a considerable period of hours doing it, or seeking it if they don’ t have it. Thus, it seems likely that, *a priori*, work should be a major factor of people’ s utility or happiness. Well–being at work can broadly be defined as the overall quality of an employee’ s experience at work (Warr, 1987).

Well–being is a multidimensional concept, and therefore, there are different conceptualisations of well–being in the literature (Van De Voorde et al., 2012). These can broadly be classified into *hedonic* and *eudemonic* measures of well–being (Bryson et al., 2015). Hedonic measures of well–being focuses on the type of effective feelings that a person experiences in his/her job (for example, contentment). Whereas, eudemonic approaches to well–being focus on the extent to which a person experience feelings that are considered to demonstrate *good mental health*, for example, the extent to which they feel a sense of purpose in their job.

Given different dimensions and conceptualizations of well–being, employees’ job satisfaction therefore, is only a narrower or sub–set measure of employee well–being; it only covers those hedonic aspects of well–being or happiness that is related to the job as opposed to eudemonic aspects. According to Spector (1997), job satisfaction is simply how individuals feel about their jobs and different aspects of their jobs. Moreover, research in this area has tended to give most attention and focus to measures of employees’ work or job satisfaction (Bryson et al., 2015)

There are therefore important differences between satisfaction, happiness and well–being. For the sake of ease, however, this paper use terms such as well–being or satisfaction interchangeably. These terms, for the same reason and motivation, are also used interchangeably in prior studies by Bryson et al. (2015), Boö ckerman and Ilmakunnas (2012), De Neve and Ward (2017).

### 2.2 Theoretical Motivation: Why Might Staff Well–Being Lead to Lower Delayed Days

Healthcare incentives can either be financial or non–financial but has two main goals: 1) to motivate and raise the morale of employees to continue to perform better and, 2) have a long lasting effect on their performance (Berdud et al., 2016). An adequately designed incentive programme is an important step towards improving employee happiness and encouraging productivity. Although financial incentives are important, an extensive number of studies from behavioural economics establishes that agents respond to non–monetary motivations (Camerer et al., 2011). Therefore, non–financial incentives may be equally important. In an environment where high performers are not rewarded or talent is not managed, employees may suffer from lower job satisfaction, high burnout and therefore exert low effort. Or, in an environment where there is a lack of a system of appreciation, autonomy or opportunities to develop, may lead staff to shirk and, implementation of management practices may not produce the desired organisational benefits. In this regard, employee well–being or satisfaction could be interpreted as measure of organisations incentive systems. This implies that, healthcare managers, by adopting well–designed incentive systems can create an environment that leads to higher staff satisfaction, which in turn can positively contribute to quality of care and productivity.

Healthcare productivity has many different dimensions or manifestations. One of them is lower delayed patient transfers or discharge. There are various theoretical channels emanating from sociology and economics literature through which, higher staff satisfaction can lead to higher individual and organisational productivity in healthcare, and therefore in the current context lower hospital discharge delays. These various theoretical channels suggest that, higher staff well–being can lead to higher on–the job performance and productivity in terms of clinical and hospital activities, thus leading to higher healthcare performance such as lower delays. Higher staff well–being through such channels or mechanisms for example, may lead to better patient flow, discharge planning, reducing errors and waste, speedier and better assessments and faster turnaround of diagnostic times.

The first theoretical channel is through increased motivation. Human capital theories from sociology and management (Herzberg et al. 2011, Maslow 1943 and MacGregor 1960) view labour as key organisational assets, rather than expandable communities that create substantial value for the organisation through innovation or building client relationships. In Zingales (2000), for example, it is human and not physical capital that is regarded as the main asset in many firms for quality and innovation. These theories argue that, higher levels of employee well–being lead employees to identify themselves with the organisation and its goals, and therefore exert higher effort in the workplace. In summary, these theories argue that employee satisfaction can improve motivation that eventually can benefit healthcare organisations in terms of higher productivity. Akerlof and Yellen (1986) posits the efficiency wage theory under which increased satisfaction can induce higher effort, because an employee does not want to get fired from a satisfying job (Shapiro and Stiglitz, 1984). Or investments made by healthcare managers to raise employee satisfaction are viewed as a “gift”, and employees therefore reciprocates with a “gift” of higher effort (Akerlof, 1982; Bradler et al., 2016). Moreover, experimental studies by Bradler et al. (2016) and Oswald et al. (2015) provides evidence that higher employee well–being have a causal connection with increased productive efforts.

The second channel is through retention since high satisfaction is seen as a key recruitment tool. Employee retention is seen as a key source of value creation in service and knowledge based industries such as healthcare, pharmaceuticals and software. Experienced employees with detailed knowledge of the organisation, resources and production processes are likely to configure better solutions and make speedier decisions, whichcan enhance productivity and possibly lower delayed transfers. Theoretically, higher productivity and lower delayed transfers go hand in hand. Increasing productivity lowers delays.

The third channel is through improved cognitive function (De Neve and Oswald, 2012). Evidence from neuroscience and cognitive science further indicates that happiness is linked to higher productivity. Subjective well–being is associated with particular neurological variations, which in turn is associated with improved cognitive abilities and skills. Such neurological mediation pathways centre on the role of positive emotions (reward) in stimulating the dopaminergic system and increasing cognitive capacity for memory tasks and attention span, which can lower errors, speed up optimal decision making and help figuring out innovative practices (Fredrickson and Branigan, 2005; Pessiglione et al., 2006; Schmitz et al., 2009; Wise, 2004). And all these are conducive to higher productivity and possibly lower delayed transfers.

Other theoretical mechanisms and empirical evidences in which higher staff happiness or subjective well–being may lead to enhanced individual and healthcare productivity at workplaces are mentioned in Bryson et al. (2015) and Chang et al. (2017). These includes better health and physiology, attitudes towards work, improved immunity and endocrine function, organizational citizenship behaviour, spill–over effects reduced turnover and absenteeism. In short, staff well-being or satisfaction can theoretically improve productivity, and an improvement in productivity can lower delayed transfers.

Given this backdrop and conceptual reasoning, the following hypothesis is tested in this paper:

### Hypothesis (H1)

*Hospitals that have higher happiness or well–being score for its staff will likely experience lower delayed transfers of care for its patients*

The relationship between well–being and delayed transfers of patients will be confounded if they are both affected by the quality of management such as senior management support. Internal hospital management or organisational practices such as technology adoption, training, flexibility, communication devices etc. will be ineffective in lowering delayed discharges if the incentive systems that influence the behaviour of healthcare professionals are not adequately designed. In other words, well–designed incentives and these practices are *complements*. That is, the marginal returns to these practices depend on whether an adequately defined incentive system is in place or not. Therefore, the following hypothesis is also tested:

### Hypothesis (H2)

*Staff well–being is independently relevant for delayed transfers of care when management quality is controlled for where the coefficient is both economically and statistically significant*.

## 3 Data

### 3.1 Delayed Transfers of Care

The delays data are for 4 years from the period 2011/12 to 2014/15 and are taken from the ‘Acute and Non–Acute Delayed Transfers of Care’ dataset, NHS England. The data is at hospital level and contains monthly information on delayed transfers of care that hospitals are required to submit as part of Community Care Act 2003. The Act covers delays among adults from English acute, mental and specialists trusts. Specialist trusts that specialise in women’ s healthcare such as gynaecology, maternity and neo–natal care are excluded from the sample. Trusts that specialise in children’ s care are also excluded—they treat relatively young patients who are unlikely to require long term care and delays for these trusts are negligible. When a delayed discharge occurs, it can be either due to the local authority or due to NHS (i.e hospitals). The relevant local authority is the council that is responsible for providing adult social care where the patient resides. There exists a formal procedure for settling disputes over cases when attribution is not reached between the two institutions concerned.

The delayed discharges are measured as the total number of bed days lost over the month due to delayed patients. The data are then aggregated over the year to reach an annual figure of days delayed. The paper measures both total number of delays (i.e. due to local authority and NHS) and those attributed to NHS only. Figure 8.1 shows the distribution of total and NHS delayed days across hospitals over the sample period.

The dataset records 10 specific reasons for delayed transfers of care: awaiting completion of (acute and non–acute) assessments, awaiting public funding, awaiting further non– acute/intermediate NHS care (for example rehabilitation services), awaiting nursing and residential home replacement or availability, awaiting care package at own home, awaiting community equipment & adaptations, patient or family choice, disputes and housing—patients not covered by NHS and Community Care Act. Figures 8.2 and 8.3 depicts these causes for total and NHS–led delays respectively. As seen from these figures, awaiting further non– acute/intermediate care remains the leading cause of delayed days followed by awaiting completion of medical assessments for total delayed days and patient/public choice for NHS delays. Not all these causes however are necessarily the ‘fault’ of NHS providers i.e. hospitals (McCoy et al., 2007). These are patient/family choice, disputes and patients not covered by the social services department funding or by the Act. This paper therefore, excludes these external causes from the analysis by subtracting them from the total and NHS delays. Their distributions given in figure 8.1 also reflects this exclusion.

To examine the impact of staff–well being on major–disaggregated causes, this paper has summed the two leading causes of delays for both total and NHS delays. Since, patient/family choice are excluded from the analysis of delays, the two major causes of total and NHS delays are therefore awaiting further non–acute/intermediate care and medical completion assessments. Their respective distributions are given in figure 8.4.

### 3.2 Staff Well–Being

Measures of NHS staff well–being are taken from the NHS Staff Survey (NSS) data for the years 2010–2014. It is constructed by summing 3 sets of questions: *job satisfaction, staff satisfaction with the quality and patient care they are able to deliver* and *recommendation of their organisation as a place to work and receive treatment*. The third set of question was used by Pinder et al. (2013) in their study of staff satisfaction and mortality performance for NHS hospitals. We can easily identify the occupation of the respondents who answered the NSS. In addition to overall measure, staff categories for clinical/medical staff and nursing staff were also included for analysis.

Staff job–satisfaction score is measured on a scale of 1–5 and is an average of 7 questions. They are Likert–type questions and are measured on a 5–point scale from “Strongly Disagree” to “Strongly Agree”. These questions are based on the following aspects of the job: recognition for good work; support from immediate managers and colleagues; freedom to choose methods of working; amount of responsibility; opportunities to use skills; and the extent to which the hospital is seen to value the work of staff. The other two remaining questions are measured by calculating their respective positive response rates (i.e % staff answering agree or strongly agree). These 3 sets of questions are standardised on a scale of 0–100 and finally average is taken to arrive at the final well–being score for the medical staff for each hospital.

The distribution of staff satisfaction scores are given in figures 8.5 and 8.6. From the figures we can notice there is presence of considerable variations in happiness scores with some section of hospitals below and above the average score.

### 3.3 Long–Term Care and other Control Variables

The four different types of NHS trusts or hospitals that are controlled for in this study are: Teaching, Foundation Trust, Specialists and Mental Health. Each hospital type is captured using dummy variables taking the values of 0 and 1. Hospitals with acute trust status is treated as the base category.

The literature has identified many potential causes for delayed days that are related to its own internal factors and services for instance Glasby et al. (2006). One of the leading causes are the lack of rehabilitation services and shortages of skilled staff. Therefore, this study has added the number of specialised staff that consists of clinical staff providing services in rehabilitation and geriatric medicine. Delayed transfers of care are particularly associated with older patients that have complex needs and geriatric medicine often purposely decelerates the process of discharge to achieve better long term results (Challis et al., 2014; Manzano-Santaella, 2010). Clinical staff providing services at these two branches of medicine obtained from NHS Workforce Statistics are summed together and standardised by the total no. of hospitals’ clinical staff to arrive at percentage based measure. We hypothesize that hospitals’ that have higher proportion of rehabilitation and geriatric care staff will have lower delayed transfers than otherwise.

The paper controls for sizes of each trust by using data on beds from “Quarterly bed availability and occupancy” submitted to the Department of Health and published by NHS England. The beds data is available for each quarter. To construct the annual data, we use the quarterly average of total available beds at each trust. Furthermore, to account for the possible non–linearity between beds and hospital delays, the following dummy categories of beds are included as controls: 200–399, 400–599, 600–799, 800–999, 1000–1499 and 1500+ beds. The base category is 0–199 beds.

To control for case–mix at each trust, we have included the percentage of patients that are male/female, aged between 60–74 years, aged 75 or more and admitted as emergencies.

As a measure of hospital quality, we have included hospital readmission rates for two age bands: patients aged 16–74 and over or equal to 75 years old. Readmission rates are risk– adjusted and measures readmissions within 28 days of discharge. The 28–day readmission rates are indirectly standardised by age, gender, method of admission, diagnoses and procedures. To reduce simultaneity bias, the readmission data are lagged by two years. Data for 2014/15 was mean imputed using the values for the years 2011/12 to 2013/14 since NHS Digital provided the readmission rates data until 2011/12. Other measures of quality for example case–adjusted mortality were not used since they were not available for all the trusts.

Differences in delayed transfers may also be due to the presence or absence of external factors such as the availability of long–term care (LTC) beds. If no bed is available at a care home, then the patient may have to remain in hospital despite being clinically fit to be discharged. Most patients have to pay or at least partly for long term care services. And so it may take longer to find a suitable care home services that patients can afford if prices are higher. To measure the accessibility of long–term care homes in the area served by a hospital Trust, we use the June 2011 data on care home beds and prices from Laing Buisson. Using ArcGIS software, we measure the number of care home beds and average price within a distance of 10 kilometres from the hospitals whose primary clients are people aged 65 years and above or with dementia. We then take the average of their available beds and prices by using taking simple straight line distances calculated using Pythagoras’ theorem. The data on care home beds and prices are only available for one year for the year 2011. This paper therefore uses the data on long term care for 2011 for the other remaining years.

### 3.4 Management Quality

Data on hospitals management quality were taken from NSS. It consists of four practices that are collected in the form of Likert type questions and serves as an indicator of support from senior management and its quality in motivating and incentivising employees. These includes decentralisation and involvement of staff in decision making, suggesting ideas for improvement of services, effective communication between senior management and staff and, staff knowing who the senior managers are. The four practices constitute some of the key elements of high–performance work system and measure hospital staff’ s perceptions of the human resource environment (Appelbaum, 2000). The final composite measure is the positive response rate (i.e respondents who answered agree or strongly agree) of these four factors.

### 3.5 Summary Statistics

Table 9.1 provides the summary statistics for the control and dependent variables across the whole sample. The total number of observations in the sample is 847. Some of the data are however are missing. Table 9.2 provides data definitions and their sources.

Figure 8.8 depicts the relationship between staff well–being score for hospitals’ medical staff and the 4 measures of delayed days. In the figure, values of delayed days were limited to 0–10000 days. Hospitals whose well–being score for its medical staff is above the mean tend to have lower median delayed discharges of patients than otherwise. The pattern is also consistent when delays are standardised by beds or bed–days.

Figure 8.7 suggests that hospitals’ experience fewer delays (relatively more dark green dots) whose well–being and management quality scores exceed their respective median (29 and 53.6 respectively). Conversely, hospitals’ whose management quality and well–being scores are below their medians experience more delays in discharging their patients i.e. relatively more lighter and non–green dots. The pattern remains when delays are standardised for example by hospital bed days.

## 4 Analytical Framework

The dependent variables are measures of count of delayed days i.e. non–negative and integer valued. *Y*_*it*_ is days of delays in hospital *i* in year *t*. The traditional approach to modelling count dependent variables is to use Poisson regression. The Poisson process assumes however that the variance equals the mean i.e. *Var*(*Y*) = *E*(*Y*) = *µ*. Empirically, we often find that data is right skewed and exhibits significant *over–dispersion* where variance is significantly larger than the mean (figures 8.1 and 8.4). Specifically:

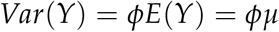

where *ϕ >* 1. The presence of over–dispersion will produce consistent estimates but the standard errors will be underestimated. An alternative approach to model count data with over– dispersion is to use Negative Binomial (NB) regressions. NB regression is a generalization of Poisson regression which loosens the restrictive assumption that variance is equal to the mean. The traditional NB model known as NB2 is based on the Poisson–gamma mixture distribution. This formulation is popular because it allows the modelling of Poisson heterogeneity using a gamma distribution. Cameron and Trivedi (2005), Cameron and Trivedi (2013), Hilbe (2011) and Winkelmann (2013) present a much more detailed and technical description of NB models and over–dispersion.

Since this paper uses a longitudinal research design, we employ the conditional fixed effects NB model derived by Hausman et al. (1984). The probability density function for hospital *i* in year *t* can be expressed as:

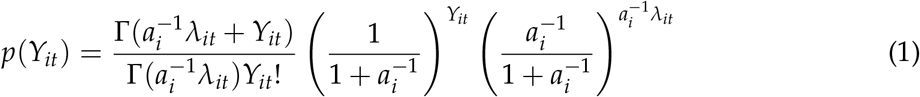

where Γ(*·*) is the gamma function, and *a*_*i*_ is the rate of over–dispersion. The expected value and variance taken from Law et al. (2011) are:

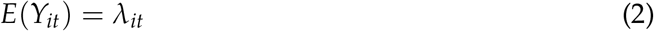

and

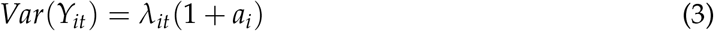

The role of the over–dispersion parameter *a*_*i*_, is to capture the heteroskedasticity in the variance estimates (Law et al., 2011). An advantage of fixed effects over pooled NB is that it allows individually different dispersion parameters and can account for heterogeneity in the data (Cameron and Trivedi, 2013; Hausman et al., 1984; Kis-Katos et al., 2014; Law et al., 2011). The random effects model assumes that the inverse of the over–distribution parameter is distributed as a beta distribution. This approach assumes that hospital specific effects is part of the error term and it is preferred when the sample is drawn from a population (Baltagi, 2008; Hsiao, 2014). However, the assumption could be violated due to the problem of endogeneity leading to biased estimates. Fixed effects on the other hand is less restrictive and allows arbitrary correlation between hospital specific intercepts and the explanatory variables.

The conditional fixed effects negative binomial model has been used recently before in academic research in many areas including economics. For instance in Law et al. (2011), Guo et al. (2014), Kibris and Metternich (2016), Schmitz (2011), Moser and Nicholas (2013), Aghion et al. (2013) and Bloom et al. (2013).

To allow meaningful comparisons in delayed days across the hospitals, we normalize the dependent variables by specifying an offset variable. We specify the offset variable as the logarithm of size measured by number of beds. In this way, the coefficients of the independent variables can be interpreted as effects on rates rather than counts where rate is defined as the number of delayed events over size. The use of exposure is superior in many ways compared to modelling rates as response variables because it makes better use of the probability distribution. The empirical model is specified as follows:

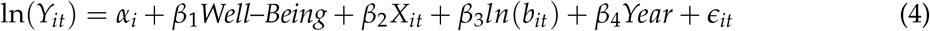

*Y*_*it*_ is the expected number of days of delays for hospital *i* in year and *t, Year* are a set of year dummies and *X*_*it*_ represents set of covariates and controls. The year dummies are included to control for year–specific systematic effects common to all hospitals that affect delayed days and for cross–sectional dependence. *b*_*it*_ is the number of beds in the hospital where we use it as an exposure term with *β*_3_ = 1. Equation (4.4.4) could also be re–written as:

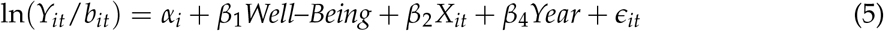

### 4.1 Econometric Issues

A important source of bias that can arise in this data is due to potential endogeneity of the staff well–being variable. Staff well–being could be correlated with unobservable hospital heterogeneity effects which could also influence mean delayed days. These unobservable factors may comprise of labour amenities, working conditions, resource constraints, management practices, institutional and organisational factors (for example, availability of intensive care unit) that may lead to both lower discharge rates and higher staff happiness. To mitigate this endogeneity problem, the paper employs a panel research design for count data based on Hausman et al. (1984) where we follow each hospital for the years 2011/12–2014/15. By using a fixed effects specification, we can eliminate the unobserved heterogeneity by conditioning out their effects (Hausman et al., 1984; Hilbe, 2011).

In addition to this and to add further support to the exogeneity assumption, this study lags the main staff well–being variable by one year. Lagging of variables is a popular method used by academic researchers in economics for example Edmans (2011) and Boö ckerman and Ilmakunnas (2012) in order to lessen the endogeneity bias in the data. Lagging variables also ensures that any potential relationship between the dependent and the independent variables is not driven by contemporaneous correlation.

A second potential problem that involves biased results is due to reverse causality. Reverse causality could easily arise in studies between staff satisfaction and organisational performance where good performance could lead to higher job–satisfaction (Boö ckerman and Il- makunnas, 2012; Edmans, 2011; Huang et al., 2015). However, we expect that delayed transfers of care will cause fewer reverse causality issues than hospital mortality, profits/losses or waiting times. Outcome measures such as mortality rates, financial indicators and waiting times are relatively popular metrics to gauge hospital performances and, are frequently used by the government to design their policies. Moreover, to further attenuate the reverse causality bias, we lag the staff well–being variable as suggested in Boö ckerman and Ilmakunnas (2012).

Allison and Waterman (2002), Greene (2007) and Guimaraes (2008) however pointed that the conditional fixed effects NB model by Hausman et al. (1984) is not a “true” fixed effects model in the sense that it does not control for all unobserved time–invariant characteristics. It models individually different dispersion parameters (which affects mean and variances) but unlike traditional fixed effects model, it allows the estimation of time–invariant variables for instance teaching status (Cameron and Trivedi, 2005). Guimaraes (2008) pointed out that the hospital fixed effects will be removed only if there is a specific functional relationship between the hospital fixed effects and the dispersion parameter. However, this assumption cannot be verified because the code of the test developed by the author has not been completely examined. In addition, the test does not work when an offset variable is included (Law et al., 2011). An alternative is to implement unconditional fixed effects using hospital dummies. There are two problems with this approach however. Firstly, there is an issue of computational complexity. Secondly, implementing this procedure especially with a short panel may cause incidental parameter bias problem.

A second alternative is to use the correlated random effects approach of Mundlak (1978) and Chamberlain (1982) and explained further in Wooldridge (2013). This approach also applies to non–liner panels and involves adding averages of time varying variables and their corresponding variables in level form (Wooldridge, 2009). The variables in level forms are interpreted as the fixed effects coefficients. This approach is tested using the Random Effects framework that allows estimation of variables that are either time–invariant and/or with minimal within–cluster variation. It therefore, provides a suitable alternative to Fixed Effects estimation. In addition to the conditional Fixed Effects of Hausman et al. (1984), this alternative approach is used for further robustness of the results.

## 5 Analysis

The over–dispersion test as explained in Hilbe (2011) indicates significant Poisson dispersion for the four delayed dependent variables; the Pearson dispersion statistics are considerably greater than 1. This therefore supports the use of negative binomial regression model.

Table 9.3 reports the first set of results involving total delayed transfers of care and consists of 6 models. All the models and the subsequent analyses were estimated using conditional fixed effects negative binomial regression to account for unobserved hospital heterogeneity. Hospital beds as an exposure term is added with a coefficient equal to 1. This in effect standardizes delays by bed. Categories of hospitals’ bed size are added to capture economies and diseconomies of scale. Finally, all the models include year dummies to control for temporal variation and year fixed effects. The coefficients are interpreted as proportionate change in days of delays from one–unit increase in explanatory variables.

Model 1 consists of basic hospital and patient level variables. Models 2, 3 and 4 adds hospital quality, long term care and staff variables respectively. The primary variable of interest, staff satisfaction is added in model 5. As in Pinder et al. (2013), staff satisfaction is added for two different categories of staff: nursing and medical. The two staff satisfaction variables were lagged by one–year and were included to examine whether there exists any heterogeneity in their effects. The inclusion of staff satisfaction variables are supported by the in–sample fit statistics provided by the AIC and BIC results.

Consistent with prior theoretical predictions, staff satisfaction is significantly associated with hospital delayed discharges with the correct expected negative coefficient. However, the relationship is significant only for clinical staffs’ well–being score. The relationship is robust to the inclusion of hospitals management quality score in model 6. The AIC and BIC results indicates a small improvement but still significant. According to Burnham and Anderson (2003), differences in the values of AIC of around 4 to 7 corresponds to roughly 95% significance. The analysis in table 9.3 suggests that controlling for hospital, long–term care and other factors, an increase in medical staff satisfaction score by 1–unit is associated with 0.37% or 0.4% (rounded to 3 decimal places) lower delayed discharge rates (model 5). In terms of Incidence Rate Ratios (IRR) which is achieved by exponentiating the estimated coefficient, an one–unit increase in staff satisfaction score is associated with lower rates of delayed days by the multiple of *e*^−0.004^≈ 0.996. When management quality is included in model 6, staff satisfaction is associated with 0.36% or 0.4% (rounded to 3 decimal places) lower rates of delays. In both the models, well–being score for medical staff appears significant at 5% whereas for nursing staff it is insignificant.

Table 9.4 reports the results for NHS led delays i.e. delays that are associated with hospitals only. Similar to 9.3, model 5 of Table 9.4 indicates that medical staff satisfaction scores are associated with lower hospital discharge rates for its patients and is robust to the inclusion of management quality in model 6. In model 5, controlling for hospital, patient, long–term care and other factors, an 1–unit increase in medical well–being score is associated with 0.44% or 0.4% (rounded to 3 decimal places) lower hospital delayed days per bed. Whereas in model 6, a unit increase is associated with 0.43% or 0.4% (rounded to 3 decimal places) delays. Well– being scores for nursing staff in models 5 and 6 has the expected negative sign but fails to appear significant in either models.

Tables 9.5 and 9.6 report the results for major and major–NHS causes of hospital delays. The results again indicate that well–being scores for hospitals’ clinical staff are associated with lower delays. For major causes of delayed transfers, clinical staff well–being is associated with 0.4% (to 3 decimal places) lower delays whereas it is associated with 0.5% (3 decimal places) lower delays for NHS related major causes in 9.6 and is significant at 5% level.

In summary, the results suggest that adequate incentive systems and staff well–being play a critical and statistically significant role in alleviating delayed hospital discharges and therefore in raising productivity. In particular, the current findings imply that the clinical staff play a key role in effective planning and discharging of patients and therefore maintaining efficient patient flow. A first–order auto regression of clinical staff satisfaction variable reveals an autoregressive coefficient of order -0.18 using fixed effects and is significant at less than 1%. This implies that on average, staff satisfaction scores for hospitals’ clinical staff has decreased over the sample period and is consistent with the rising trend in hospital delayed transfers of care in figure 1. The results in Tables 9.3 to 9.6 indicate that hospital management quality is effective in reducing hospital discharge rates at acceptable levels of statistical significance. For instance in model 6 of table 9.4, an unit increase in management quality score is associated with 1.1% lower NHS–hospital delayed days.

Moreover, the results further indicate that in addition to management quality, staff well– being is independently effective in reducing delayed transfers of care and therefore, highlights the importance of an well designed incentive systems.

Amongst the other covariates in Tables 9.3 to 9.6, hospitals with Foundation Trust statuses and availability of long–term care beds are associated with lower delays. For instance in table 9.3, hospitals that have Foundation Trust statuses are associated with 45%–25% lower delays controlling for other factors. Since long term care beds is measured using natural logs, the coefficient has a percentage interpretation: an 1% increase in the availability of long term care beds is associated with 0.21%–0.29% lower delays (table 9.3). Amongst patient factors, percentage of patients treated in the 60–74 age group are associated with lower delays. Whereas, percentage of patients in the age group 75 years and above and male patients are associated with higher hospital delays. For instance in table 9.4, hospitals’ that treat a greater percentage of patients in the 60–74 age group are associated with 0.040%–0.038% lower delays (models 3 to 6). In Table 9.6 on the other hand, treating 1% more patients in the age category 75+ is associated with 0.036%–0.026% higher delays. The year dummies are generally significant especially for the year 2014/15 where there has been a sharp increase in delayed transfers of care in the NHS.

Empirical results so far do not support the effectiveness of well–being for the nursing staff in alleviating delayed discharges. As a robustness exercise, the analysis in Tables 9.3 to 9.6 were repeated using only nursing staff satisfaction and then the equivalent using only total staff (i.e including both clinical, non–clinical staff and other staff). However, the data and resultant analysis finds no evidence of their significance. The lack of evidence for satisfaction of hospitals’ total staff may be due to the presence of large proportion of non–medical staff that are not directly involved in organising and delivering patient care under both acute and non– acute care settings. Staff responses from non–clinical groups or from less acute care settings may also create randomness and statistical noise.

Table 9.7 report the result for the four delayed variables using only the clinical satisfaction scores. Due to convergence issues, model 3 for major delayed days was estimated using finished consultant episode bed days as the exposure term rather than hospital beds. Variable bed–days and hospital beds are highly significant with correlation coefficient of around 0.95. Table 9.8 repeats the findings in Table 9.7 by excluding specialist trust hospitals. The motivation for excluding specialist trust is derived from the fact that only a portion of hospital trusts are specialists i.e. 6% and, they tend to have a different cost structure compared to other acute trusts. And, specialist trusts also have the highest mean staff satisfaction scores. To ensure that the current results are not driven by a small sample of hospitals, specialists are excluded in table 9.8. Due to convergence issues, models 3 and 4 for NHS delays was estimated using bed– days as the exposure term. As in the above findings, well–being score of hospitals clinical staff appears statistically significant at 1% or 5% significance levels where higher satisfaction scores are associated with fewer delays. The effects are stronger for Total and NHS delays (models 1 and 2) with an estimated coefficient of 0.5% (rounded to 3 decimal places).

### 5.1 Robustness Tests

Although the conditional Fixed Effects negative binomial regression remains frequently used, it has been criticised because the conditional Fixed Effects is not a ‘true’ fixed–effects model as it does not completely condition out the fixed effects, or more precisely, only given restrictive assumptions. As a robustness exercise, the paper adopts the approach of Mundlak (1978) and Chamberlain (1982) that provide an alternative to fixed effects and also allows estimation of time–invariant covariates. This approach, known as correlated random effects (CRE) involves adding the averages of time–varying explanatory variables in the model along with their associated variables in level form. The coefficient of variables in level form provide the fixed effects estimates. As in Goerke and Pannenberg (2011) we only take the means of some time–varying explanatory variables namely staff satisfaction and hospitals’ management quality. Most of the time–varying variables for example patient factors are slow–moving and have minimal within–cluster variations. Therefore, fixed effects estimates of such variables will be inefficient as they will be estimated with high standard errors.

The results of CRE approach are reported in tables 9.9 and 9.10 for the full and without the specialist trusts sample respectively. The medical staff satisfaction variables are lagged by one year to minimize the impact of endogenous bias and to ensure that results are not driven my mere correlation. In both the tables, the fixed–effects estimates for the medical staff satisfaction variables appears significant at all acceptable levels of statistical significance with the expected correct sign. So, although the conditional method does not allow for ‘true’ fixed effects, the above estimates are about the same, suggesting that the bias in the conditional method is minimal with the current data. Due to model 4 for major NHS led delays not being able to converge, it was estimated using bed–days as the exposure term in tables 9.9 and 9.10 and, using a single readmission rate variable (readmission rate variable for patients aged 16 years and above) in table 9.9.

In columns 1 and 2 of tables 9.9 and 9.10 for total and NHS delays, a unit increase in medical staff satisfaction score is associated with 0.4% less delays holding all other factors constant. The coefficient is higher for major NHS led causes of delays with coefficients of 0.43% and 0.46% for total and without specialist hospital samples respectively and are significant at 5% level. Hospital senior management quality variable also appears statistically significant with the expected negative signs.

For further robustness checks, the CRE approach in tables 9.9 and 9.10 was implemented with the averages of all the time varying explanatory variables and also including nursing staff satisfaction. And for the sake of completeness, Tables 9.3 to 9.6 was also estimated using the CRE formulation. The results are robust to all these alternative specifications and to inclusion of hospital interaction terms. The results are also robust using a hybrid approach when variables in level form are instead replaced by their demeaned version as suggested in Allison (2005, Chapter 4.4).

The current measure of delayed days excludes external causes of delays like patient or family choice. The results and conclusions however in general remains the same even when these external reasons for delays are not excluded. The results are also consistent when considering a single readmission rate variable for ages 16 and above.

To test the potential problem of reverse causation, future *(t + 1)* values of medical staff satisfaction was regressed on current *(t)* values of delayed days along with hospital covariates such as teaching status, foundation trust and hospital staff. Both fixed and random effects were implemented along with Hausman and Mundlak test. The tests overwhelmingly indicate fixed effects as the preferred estimation procedure over random effects. The results further suggests no evidence of reverse causation as the coefficient for delayed days was largely insignificant.

### 5.2 Test for Exogeneity

To test the exogeneity of the medical staff satisfaction variable, the paper adopts a test that was suggested recently by Wooldridge (2015). The test provides a robust way to test the exogeneity of potential endogenous variable in non–linear models. The approach is divided into two stages. In the first stage, the reduced form of endogenous regressor is regressed on instrumental variable(s) by using Poisson fixed effects. The estimated fixed effect residuals in the first stage is then added to fixed effects Poisson estimation in the second stage. A robust Wald test is then used to test the significance of the residuals where the null hypothesis assumes exogeneity.

The chosen instrument measures the “fairness and effectiveness of incident reporting procedures” in hospitals and was obtained from NSS. The instrument is correlated with staff satisfaction variable conditional on other covariates (i.e first–stage exists) and, is assumed to be uncorrelated with the error term conditional on other covariates (exclusion restriction). The results of the second stage regression are reported in table 9.11. The estimated fixed effects residuals are very close to zero and insignificant across all the four measures of delays. Therefore, the robust Wald’ s test cannot reject the null hypothesis for exogeneity of the medical staff satisfaction variable.

The medical staff satisfaction variable used for the exogeneity test are its current values. However, the general conclusion remains the same if its lagged values are taken instead.

## 6 Discussion

The principal variable of interest, staff well–being is consistently associated with lower delayed days or discharge rates and therefore higher productivity. The relationship is statistically significant for clinical staff category of hospitals’ workforce. Prior studies have found both nursing and clinical staff well–being are associated with better hospital performances such as mortality. The current data and the analysis however does not find significant association between nursing or total staff in relation to lower hospital delayed discharges.

The findings are robust to patient, long–term care and other controls, alternative measures of delays and to other robustness checks and specifications. Staff well–being is also robust to the inclusion of hospitals’ management quality score which was introduced as an additional predictor to prevent any confounding relationships. Estimated coefficient for a unit increase in staff happiness or well–being score is associated with 0.40%–0.50% lower delayed transfers of care standardizing for hospital beds, depending on the specification and measure used. In terms of IRR, a unit increase in well–being score is associated with lower rates of delayed days by a multiple of 0.995 to 0.996, holding other factors fixed. The findings imply that as much as the importance that is placed on external factors such as availability of care home beds or social care expenditure, hospitals own internal processes and factors can play a very important role in reducing delayed discharges for its patients. The coefficient for hospitals’ staff well–being score supersedes the coefficients for case–mix and social care factors such as availability of LTC beds (0.21%–0.39%), which has recently received much attention in both academic and non–academic places in lowering delayed transfers of care (Donnelly, 2016; Gaughan et al., 2015, 2016; Manzano-Santaella, 2010). The coefficients also appears relatively larger in terms of magnitude in comparison to other hospital internal factors such as the presence of both skilled geriatric and rehabilitation staff in tables 9.9 and 9.10.

The findings suggest the importance of medical staff such as NHS consultants, junior and A&E doctors and their well–being for organisational performance. They are highly trained with high levels of human capital and remain a predominant driving force and leaders for providing and controlling patient’ s use of healthcare services such as hospital beds. The findings are consistent with the conceptual framework where medical staff are seen as central figures for hospital processes in maintaining efficient patient flow and throughput. And, identifying and helping to remove “bottlenecks” in the system thereby ensuring beds become available when needed. In a recent study involving delays in paediatric care units, physician led–delays such as discharge planning, paperwork and prioritisation during medical rounds are identified as the major barriers in ensuring timely discharges of its patients (Mustafa and Mahgoub, 2016). The role of medical staff in reducing delayed days also becomes more evident from the fact that some of the major reasons for delays are either due to patients awaiting further non–acute or intermediate care and completion of medical and non–medical assessments. The empirical findings of this paper are also consistent with the theoretical predictions of the human capital theories, Akerlof (1982) and happy–productive worker thesis which posits that staff satisfaction causes stronger organizational performances through improved recruitment, retention, engagement and productivity at their workplaces. Furthermore, the general findings of this paper are also in line with previous empirical evidences of staff satisfaction for non–healthcare industries.

In addition to staff well–being, hospitals management quality appears statistically significant in reducing delays at acceptable levels of significance. Presence of specialised staff appears statically significant only in the CRE models for certain delayed days where greater availability of geriatric and rehabilitation staff are associated with lower delays. Amongst the patient factors, male and older patients are consistently associated with higher delays. This finding is consistent with intuition and with prior studies as relatively older patients are generally more complex to treat, have higher co–morbidities and have greater non–acute care needs which makes them difficult to discharge early. As in Gaughan et al. (2015) and Forder (2009), the present study finds consistent role in the availability of social services such as long– term care beds in reducing hospital delays. In general, no significant association was found between delayed discharges and hospital quality measured by readmission rates.

The study however is not without its limitations. Firstly, the study utilises a small panel sample of 4 years. Further study might look to extend the time horizon to further analyse on the robustness of the findings. Secondly, there are some data limitations for delayed transfers of care obtained from NHS England. It is not clear whether all the providers are using the definitions of delayed transfers of care and causes of delays in the same way. If not, small differences in interpretation could lead to large changes in reported delays. Thirdly, throughout this paper the terms hospital and trust were interchanged repeatedly. In NHS, a trust may have one site or hospital or, may have multiple hospitals. The staff satisfaction data is by trust level and not by hospitals. Future research could utilise the hospital based data rather than trust. Fourthly, the measure of satisfaction that is employed could be improved by employing more job–related effects and hedonic measures of employee well–being such as anxiety, depression, boredom etc. Most of these data are not collected by NSS or they were not available for the study period. The measure of satisfaction also does not compartmentalise medical staff (or nursing staff) into various titles such as NHS consultants, registrars etc. As a result, we may not be able to uncover some of the heterogeneity that there may exist in the responses. Fifthly, the current study has focused on one of many measures of hospital internal factors. A more robust and a comprehensive study would also account and analyse the impact of other key factors in understanding the drivers of hospital discharge delays. Moreover, the management variable used in this paper consists of coarse number of practices. A greater understanding would require additional data on practices to study the interaction between management and well–being and to examine whether latter is independently relevant in reducing delays. Future research could address these and other shortcomings, employ a more robust research design that addresses endogeneity with granular data and, could also incorporate objective along with subjective measures of well–being. Finally, although this paper have been able to establish an empirical link, further research in the future could attempt to shed some light on the potential *mechanisms*.

The findings have implications for both hospital managers and policy makers. Firstly, the study implies incorporating more innovative human resource and operational management practices that are conducive to hospitals’ internal environment, staff morale and their well– being. Secondly, managers and policy makers need not necessarily raise well–being through employee expenditure programs or using financial incentives such as pay–for–performance. Organisational performance and productivity can be raised by improving the design of the incentive systems that improve staff satisfaction through non–pecuniary ways and, that focuses on greater employee engagement. Therefore, managers and policymakers can place less focus on cost–cutting measures or on remuneration systems. Through such internal measures, managers or policy makers can help to alleviate delayed transfers of patients by simply “internalizing” the problem. Thirdly, staff satisfaction may also be used as an early warning systems that can help to identify or spot poor organisational performances. This could help to predict early deterioration in delayed transfers of care situation. Given that there exists substantial and persistent variations in both total and clinical staff satisfaction scores, there also exists scope for improvements.

## 7 Conclusion

Hospital delayed transfers of care or “bed–blocking” remains a significant and persistent problem facing healthcare systems worldwide and, has wide efficiency and productivity implications. A health service’ s greatest asset is its staff. It is believed that this is the first study that attempts to examine the impact of staff well–being in reducing delayed transfers of care across the hospitals. The paper finds consistent evidence that hospitals’ that have higher well– being or satisfaction score for its medical staff are associated with lower delayed discharges and therefore higher productivity. The findings are robust to alternative measures of delayed transfers and various specifications, methods and including hospital management quality as an additional covariate.

Although the results suggest a link, the methodologies does not fully test for *causality*. Neither it is claimed in this paper to have obtained a causal relationship. The paper and its analyses have taken steps to address the prime econometric concerns and, the results are consistent with previous quasi–experimental evidence and theoretical predictions that argues higher employee satisfaction causes better organisational performances. There is therefore, a *prima facie* case for managers and policy makers to invest in complementary management practices, hospital incentive systems and other policies that aim to raise staff satisfaction on the basis of likely organisational benefits. This study also suggests that improving employee well–being will not be panacea and call for more empirical studies that combine information on delayed days, hospital performance, well–being and other internal factors.

## Data Availability

Data and statistical code are available on request from the corresponding author.

## Acknowledgements

We would like to thank the participants at 2018 Oxford International Health Congress, St. Hugh’ s College, University of Oxford. We would like to thank the Alliance Manchester Business School for the financial support to carry out this study.

## 8 Appendix Figures

**Figure 8.1.**
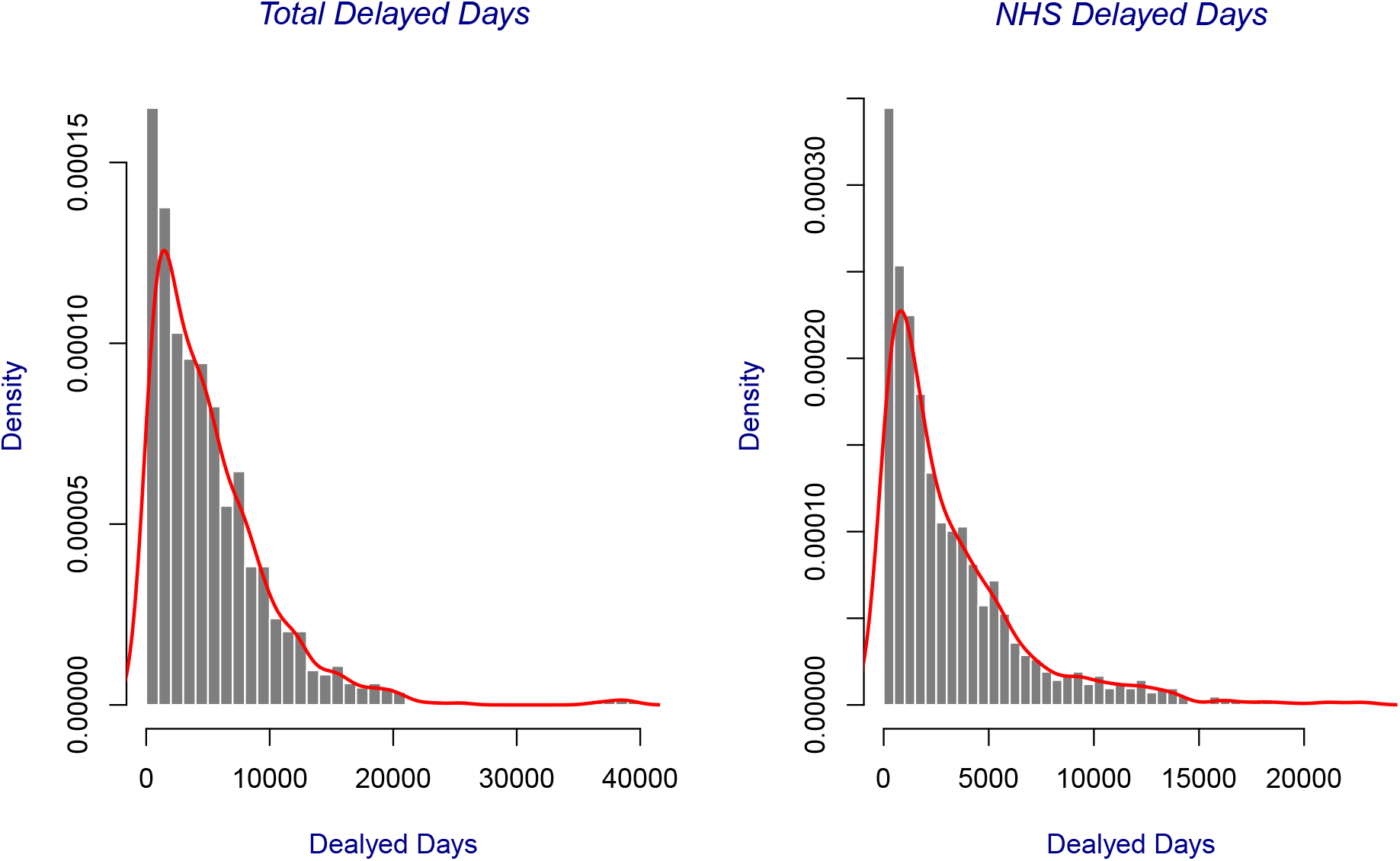
Days of Delay Distribution

**Figure 8.2.**
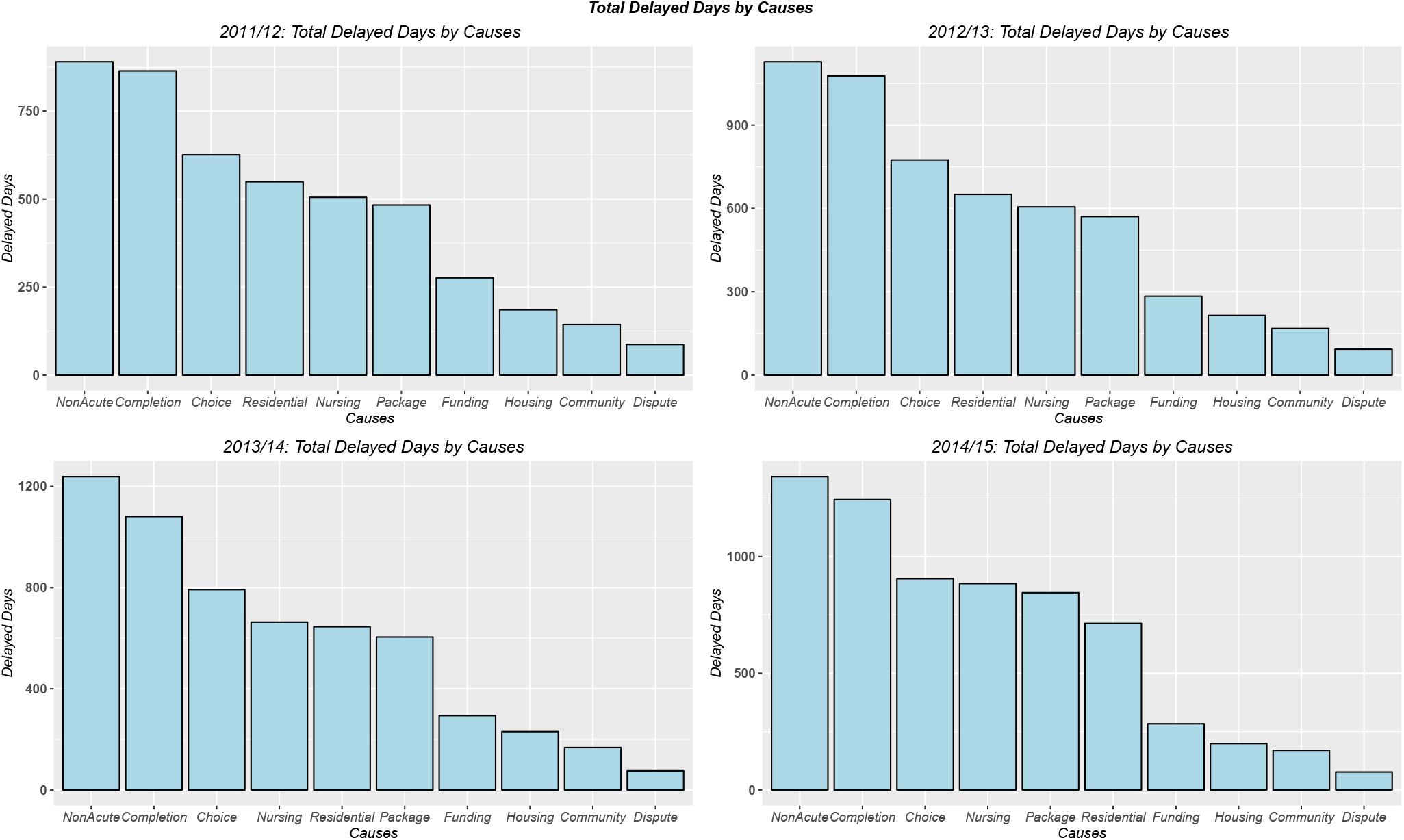
Total Days of Delays by Causes

**Figure 8.3.**
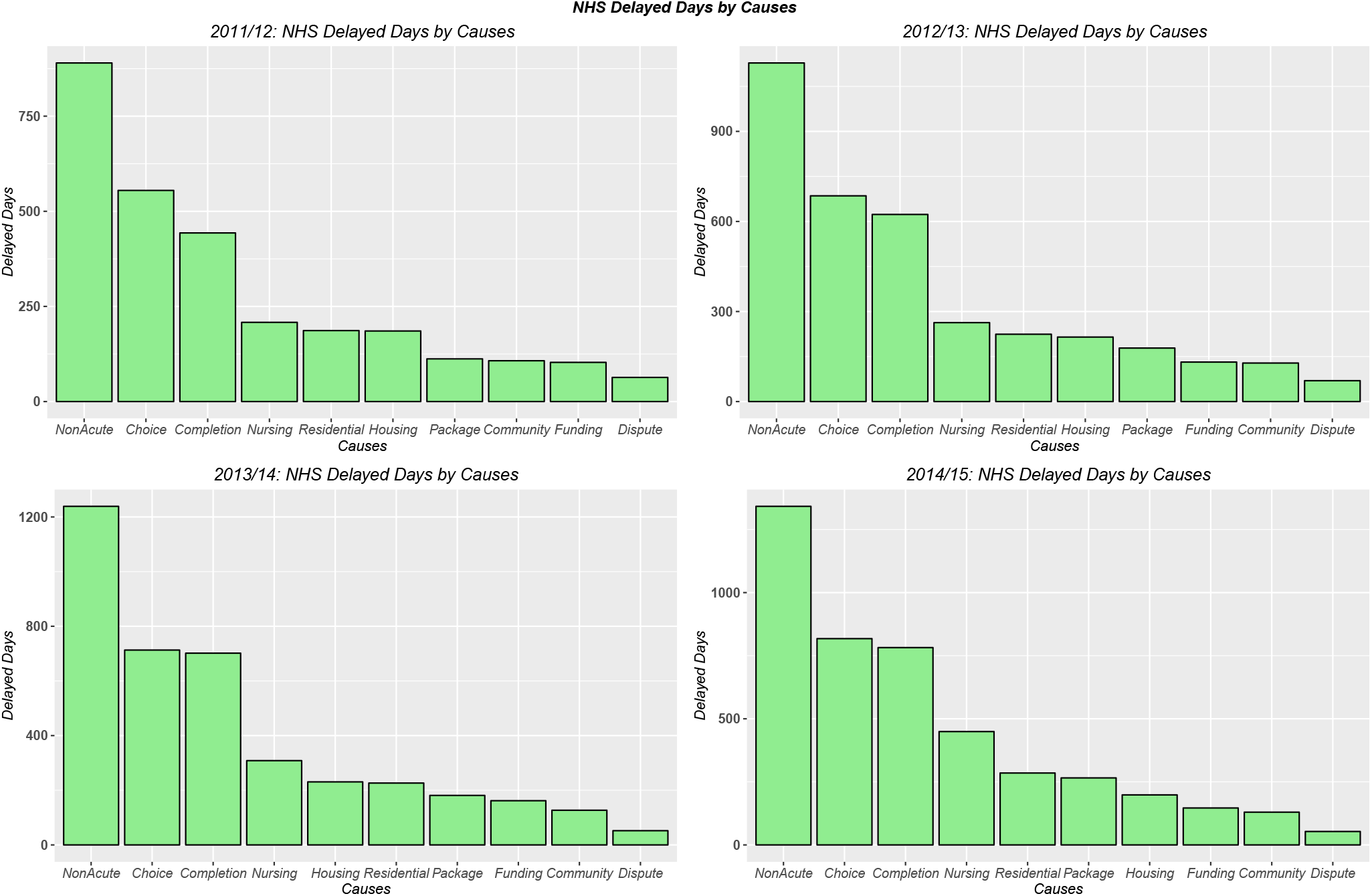
NHS Days of Delays by Causes

**Figure 8.4.**
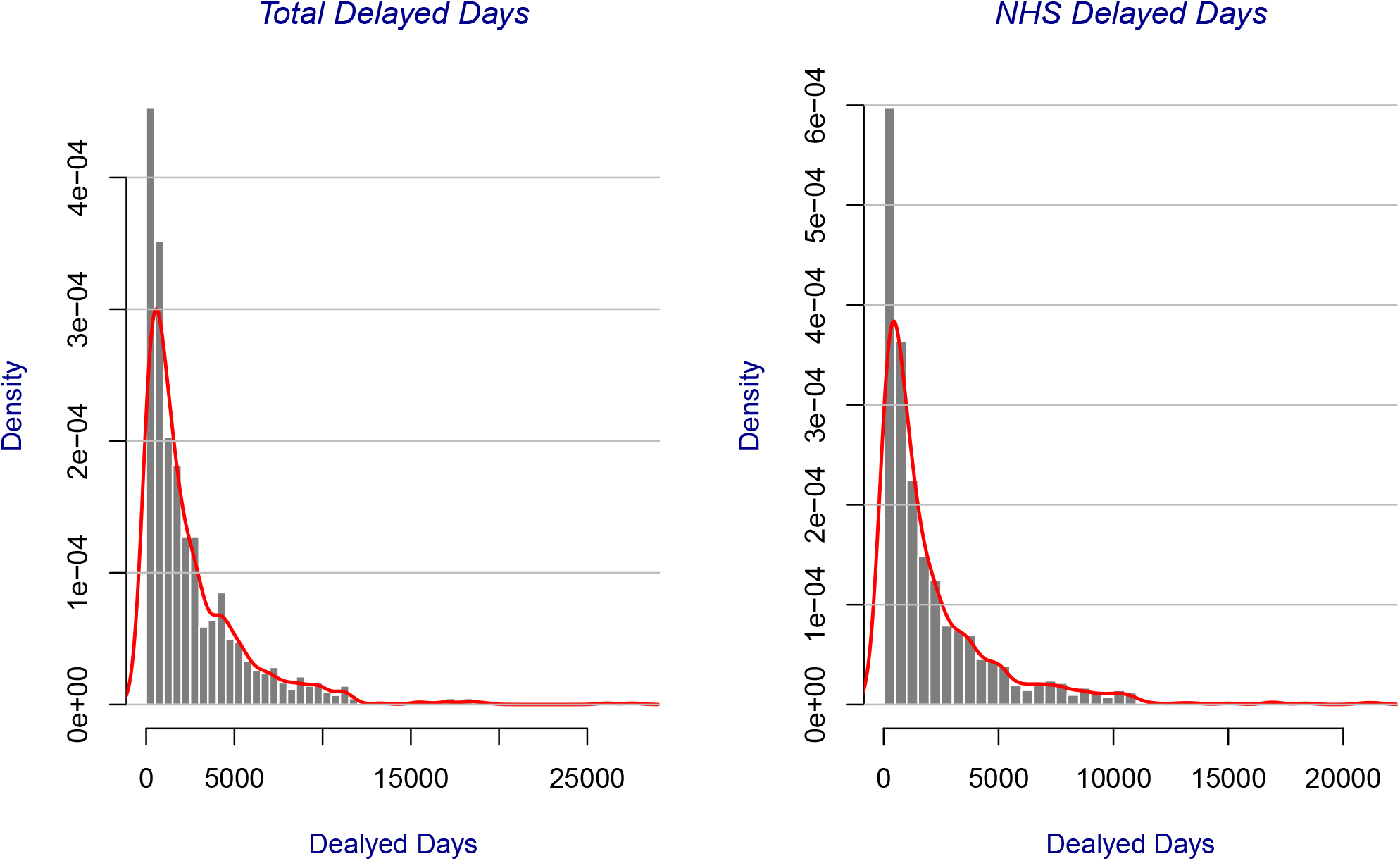
Days of Delay Distribution by Major Causes

**Figure 8.5.**
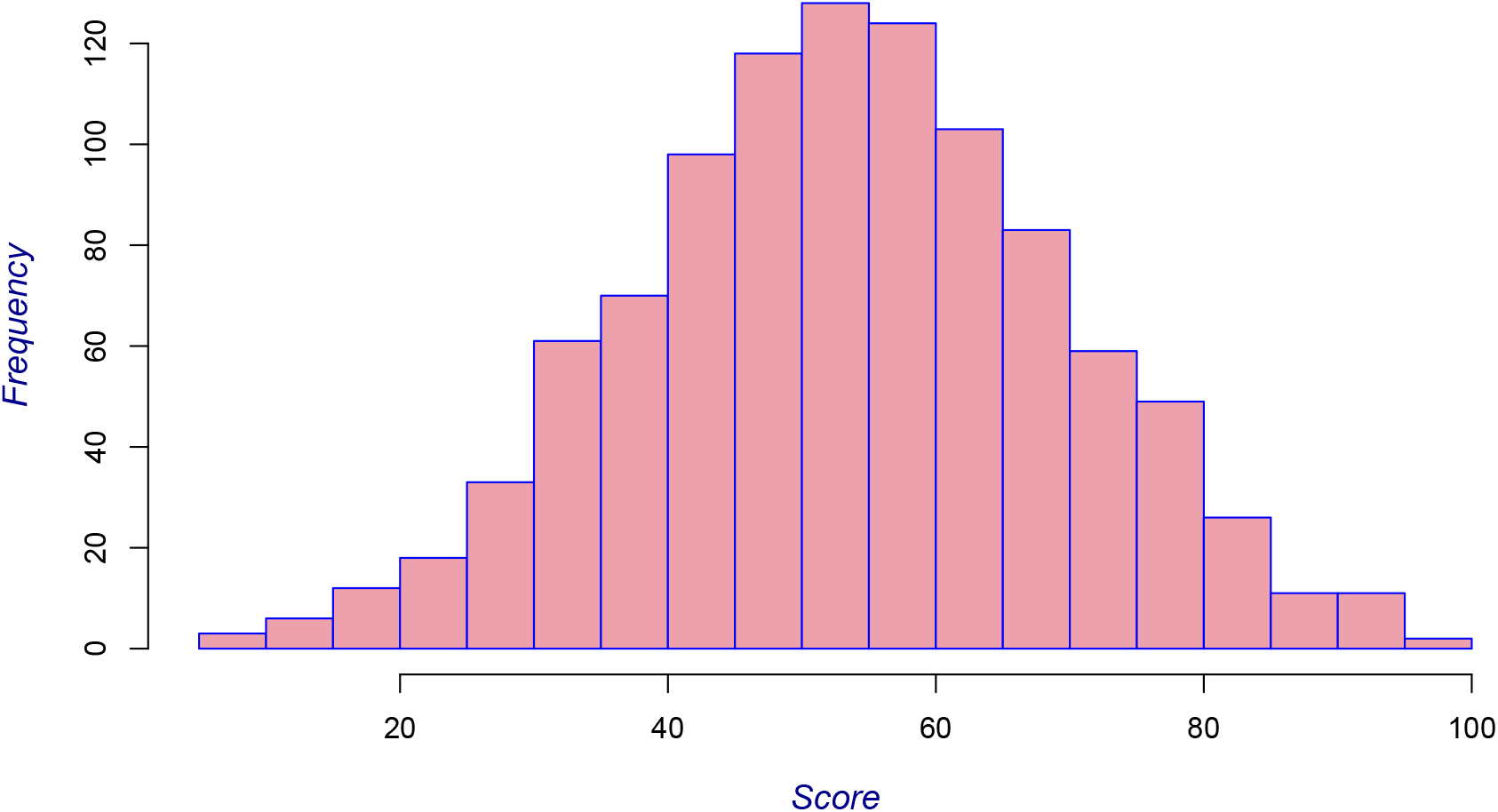
5: Histogram of Total Staff Well–Being Score

**Figure 8.6.**
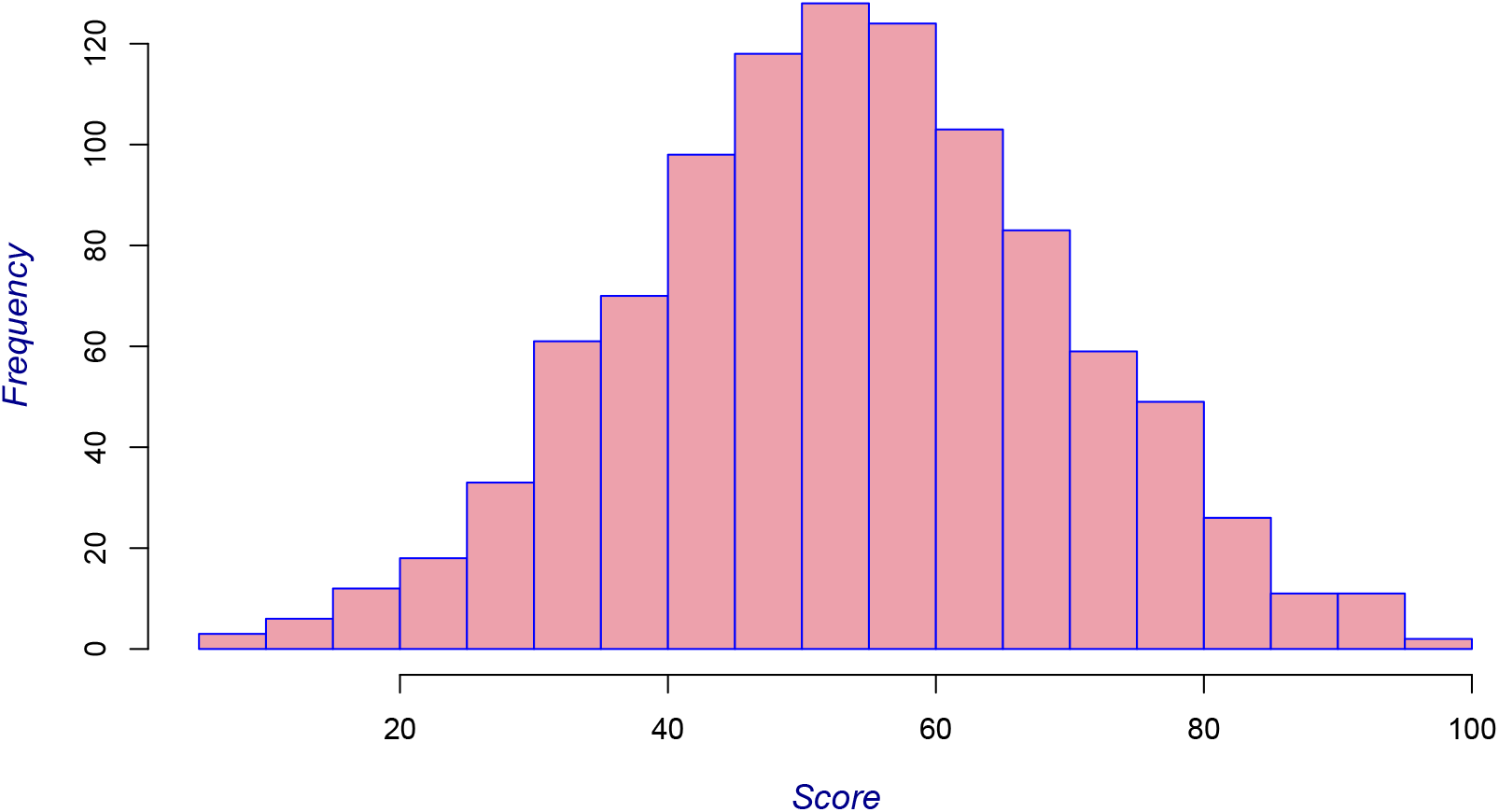
Histogram of Medical Staff Well–Being Score

**Figure 8.7.**
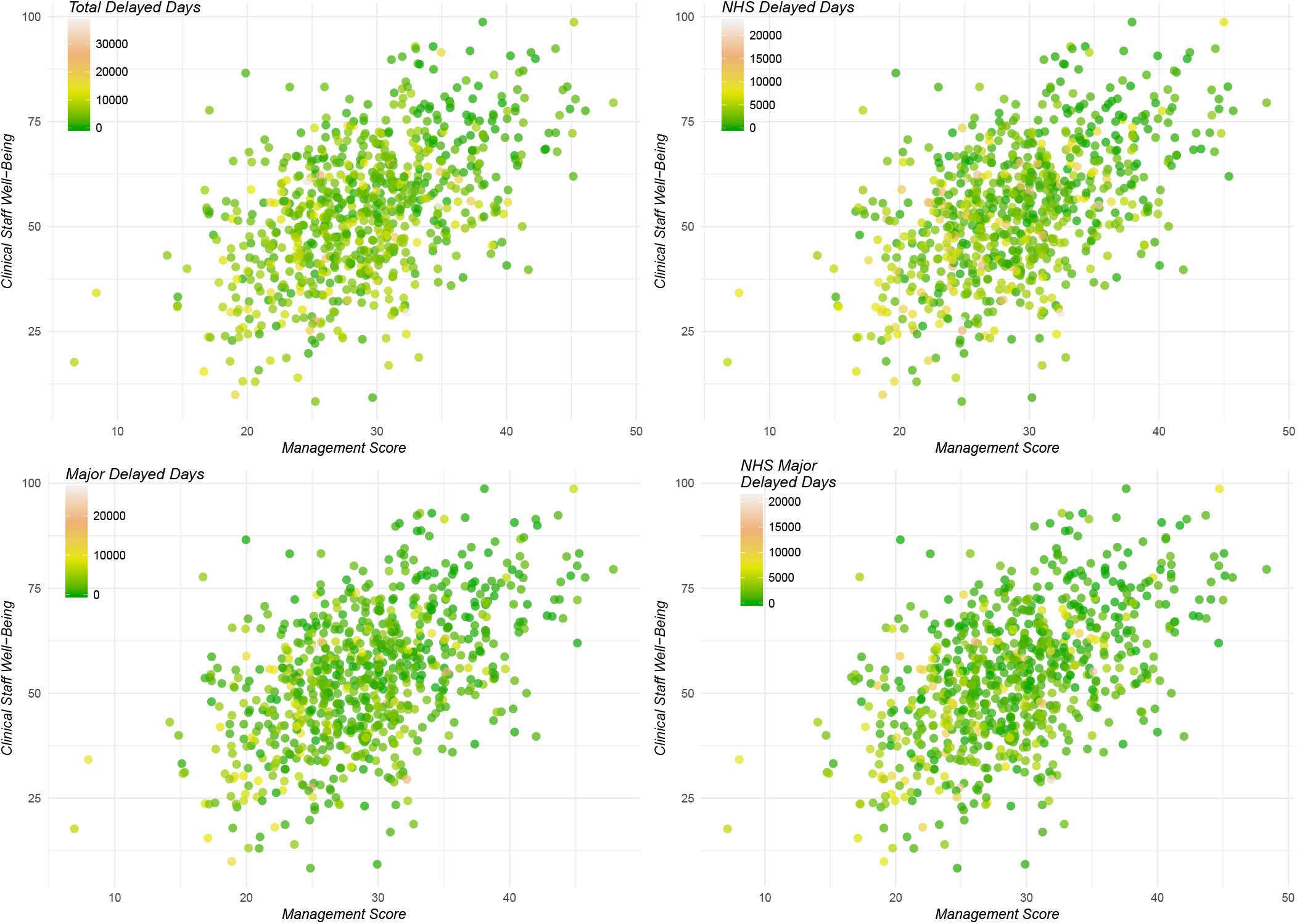
Clinical Staff Well–Being & Management

**Figure 8.8.**
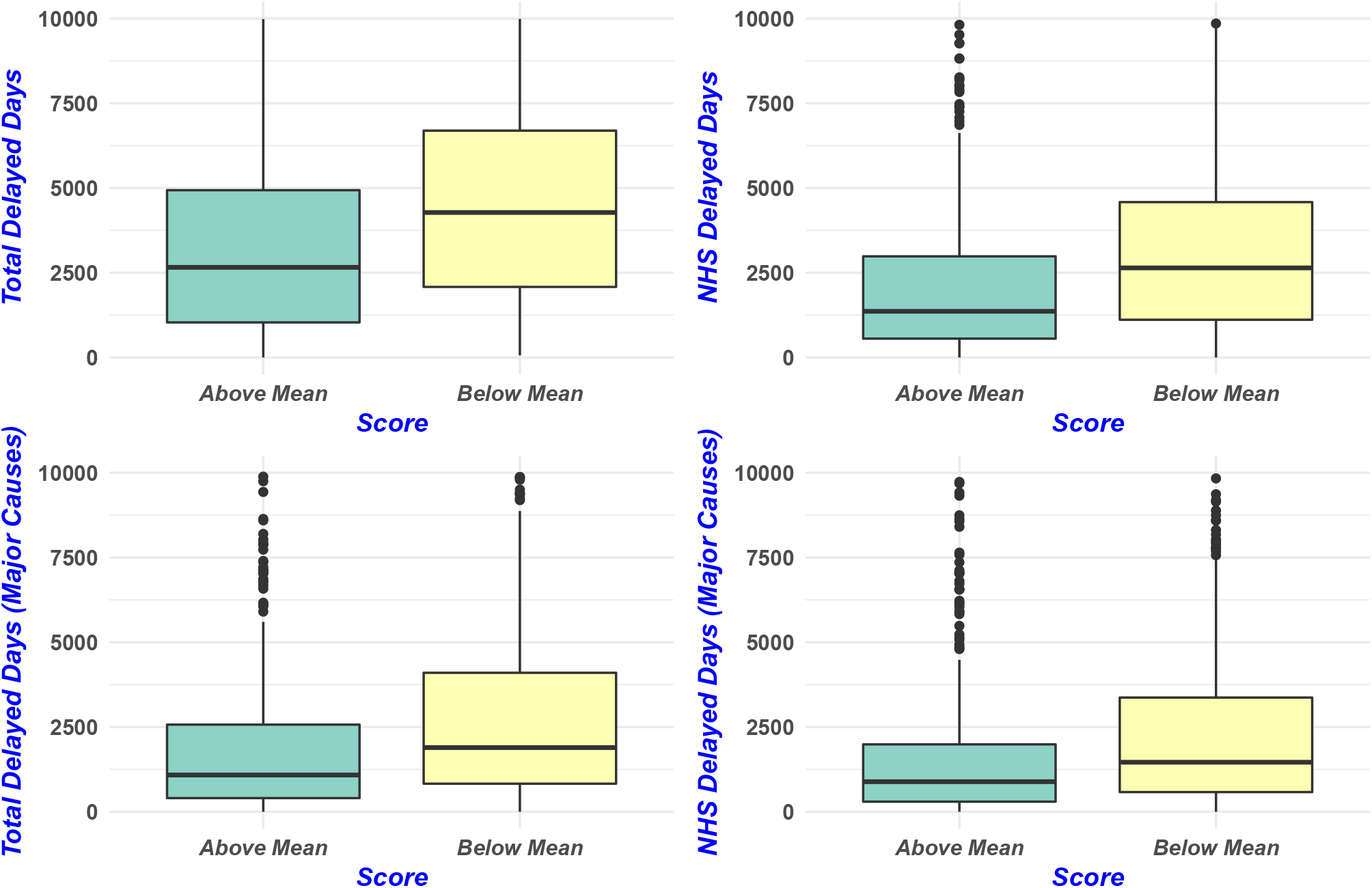
Medical Staff Satisfaction & Delayed Discharges

## 9 Appendix Tables

**Table 9.1:**
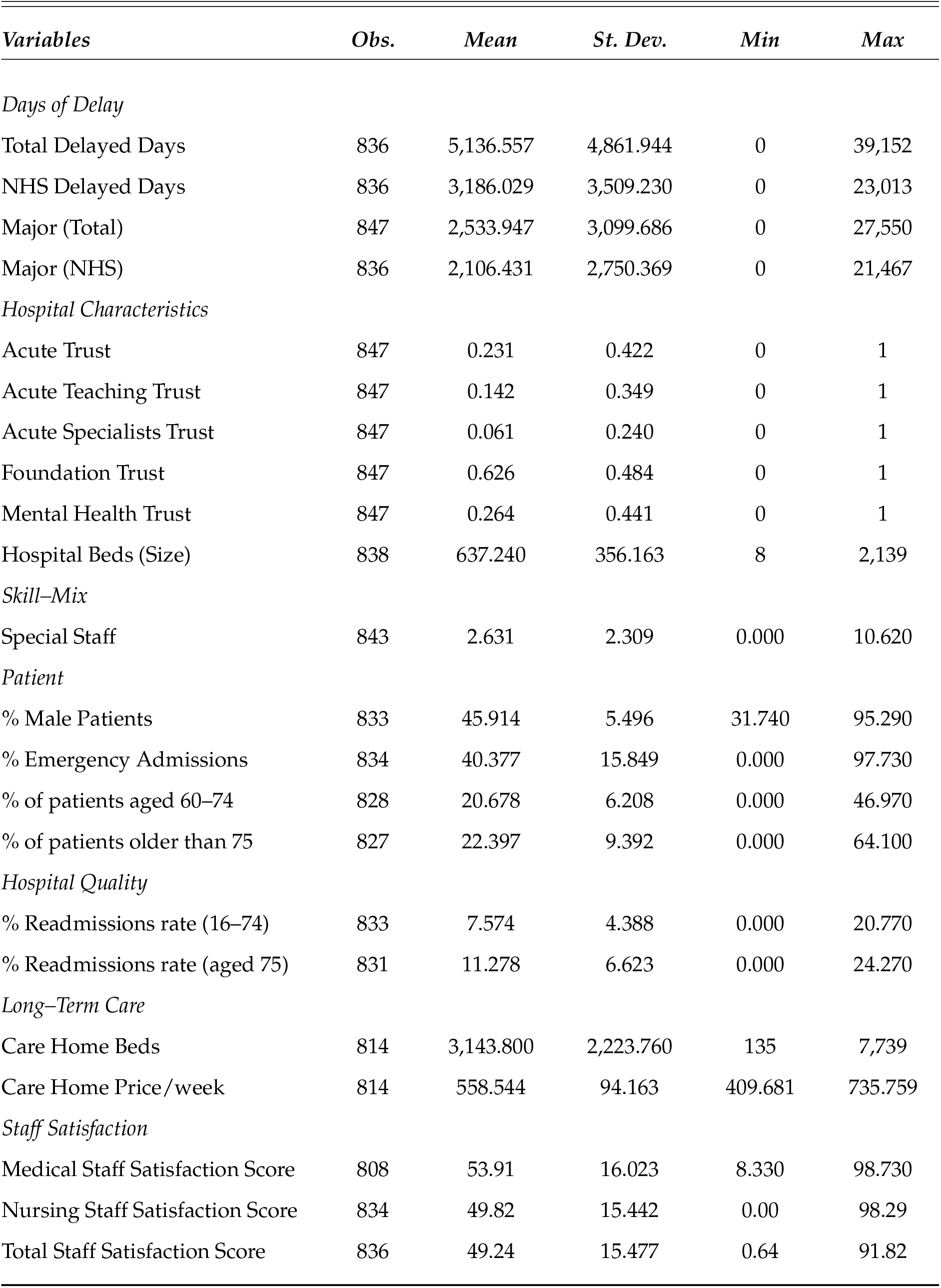
Summary Statistics

| *Variables* | *Obs.* | *Mean* | *St. Dev.* | *Min* | *Max* |
| --- | --- | --- | --- | --- | --- |
| *Days of Delay* |  |  |  |  |  |
| Total Delayed Days | 836 | 5,136.557 | 4,861.944 | 0 | 39,152 |
| NHS Delayed Days | 836 | 3,186.029 | 3,509.230 | 0 | 23,013 |
| Major (Total) | 847 | 2,533.947 | 3,099.686 | 0 | 27,550 |
| Major (NHS) | 836 | 2,106.431 | 2,750.369 | 0 | 21,467 |
| *Hospital Characteristics* |  |  |  |  |  |
| Acute Trust | 847 | 0.231 | 0.422 | 0 | 1 |
| Acute Teaching Trust | 847 | 0.142 | 0.349 | 0 | 1 |
| Acute Specialists Trust | 847 | 0.061 | 0.240 | 0 | 1 |
| Foundation Trust | 847 | 0.626 | 0.484 | 0 | 1 |
| Mental Health Trust | 847 | 0.264 | 0.441 | 0 | 1 |
| Hospital Beds (Size) | 838 | 637.240 | 356.163 | 8 | 2,139 |
| *Skill-Mix* |  |  |  |  |  |
| Special Staff | 843 | 2.631 | 2.309 | 0.000 | 10.620 |
| *Patient* |  |  |  |  |  |
| % Male Patients | 833 | 45.914 | 5.496 | 31.740 | 95.290 |
| % Emergency Admissions | 834 | 40.377 | 15.849 | 0.000 | 97.730 |
| % of patients aged 60–74 | 828 | 20.678 | 6.208 | 0.000 | 46.970 |
| % of patients older than 75 | 827 | 22.397 | 9.392 | 0.000 | 64.100 |
| *Hospital Quality* |  |  |  |  |  |
| % Readmissions rate (16–74) | 833 | 7.574 | 4.388 | 0.000 | 20.770 |
| % Readmissions rate (aged 75) | 831 | 11.278 | 6.623 | 0.000 | 24.270 |
| *Long-Term Care* |  |  |  |  |  |
| Care Home Beds | 814 | 3,143.800 | 2,223.760 | 135 | 7,739 |
| Care Home Price/week | 814 | 558.544 | 94.163 | 409.681 | 735.759 |
| *Staff Satisfaction* |  |  |  |  |  |
| Medical Staff Satisfaction Score | 808 | 53.91 | 16.023 | 8.330 | 98.730 |
| Nursing Staff Satisfaction Score | 834 | 49.82 | 15.442 | 0.00 | 98.29 |
| Total Staff Satisfaction Score | 836 | 49.24 | 15.477 | 0.64 | 91.82 |

**Table 9.2:** Data Definitions and Sources

| *Data* | *Definition* | *Source* |
| --- | --- | --- |
| ***Delayed Days*** |  |  |
| Total Delayed Days | Total annual delayed days due to both NHS and Social Care | NHS England |
| NHS Delayed Days | Total annual delayed days due to NHS | NHS England |
| Major (Total) | Delayed days due to further intermediate care and assessments (Social & NHS) | NHS England |
| Major (NHS) | Delayed days due to further intermediate care and assessments (NHS only) | NHS England |
| ***Hospital*** |  |  |
| Teaching | Equal to 1 if hospital is Teaching trust and 0 otherwise | http://www.nrls.npsa.nhs.uk/ |
| Foundation Trust | Equal to 1 if hospital has FT status and 0 otherwise | Monitor |
| Specialist Trust | Equal to 1 if specialist trust and 0 otherwise | http://www.nrls.npsa.nhs.uk/ |
| Mental Trust | Equal to 1 if mental trust and 0 otherwise |  |
| Trust Beds | Average number of available beds | NHS England |
| ***Patient*** |  |  |
| Patients aged 60–74 (%) | Proportion of patients (FCE) between 60 and 74 years of age | NHS Digital |
| Patients aged 75 (%) | Proportion of patients (FCE) aged over 74 years of age | NHS Digital |
| Proportion of Emergency patients (%) | (Emergency admissions / Total admissions) * 100 | NHS Digital |
| Proportion of Male patients (%) | (Male admissions / Total admissions) * 100 | NHS Digital |
| ***Readmission rates*** |  |  |

Table 9.2: Data Definitions and Sources
| *Data* | *Definition* | *Source* |
| --- | --- | --- |
| Readmissions Rate 16–74 (%) | Emergency hospital readmission within 28 days of discharge for patients aged 16–74 | NHS Digital |
| Readmissions Rate 75 (%) | Emergency hospital readmission within 28 days of discharge for patients aged over 74 | NHS Digital |
| *Skill–Mix* |  |  |
| Specialized/Special Staff (%) | Percentage of rehabilitation and geriatric Staff over total medical staff | NHS Workforce Statistics |
| *Long Term–Care (LTC)* |  |  |
| LTC Beds | Number of available long term care beds (2011) | LaingBuisson |
| LTC Prices | Average LTC price (2011) | LaingBuisson |
| *Staff Satisfaction* |  |  |
| Staff Well–Being/Satisfaction Score | Composite medical staff satisfaction/well–being score (0–100) | NHS Staff Surveys |

**Table 9.3:** Total Days of Delay

|  | *Model 1* | *Model 2* | *Model 3* | *Model 4* | *Model 5* | *Model 6* |
| --- | --- | --- | --- | --- | --- | --- |
| Teaching Trust | −0.4277**<br>(0.2139) | −0.4491**<br>(0.2134) | −0.2729<br>(0.2337) | −0.3527<br>(0.2395) | −0.2410<br>(0.2413) | −0.2506<br>(0.2434) |
| Foundation Trust | −0.2659**<br>(0.1107) | −0.3403***<br>(0.1134) | −0.3318***<br>(0.1152) | −0.3907***<br>(0.1155) | −0.4639***<br>(0.1120) | −0.4424***<br>(0.1130) |
| Specialist Trust | 1.4276***<br>(0.3866) | 1.3912***<br>(0.3986) | 1.5283***<br>(0.4053) | 1.4543***<br>(0.4068) | 1.1230**<br>(0.4384) | 1.0979**<br>(0.4386) |
| Mental Trust | 0.2573<br>(0.2185) | 0.4566*<br>(0.2450) | 0.4291*<br>(0.2442) | 0.3649<br>(0.2463) | −0.1005<br>(0.2848) | −0.0801<br>(0.2850) |
| % of Male Patients | 0.0286**<br>(0.0134) | 0.0221<br>(0.0140) | 0.0209<br>(0.0146) | 0.0214<br>(0.0143) | 0.0275*<br>(0.0144) | 0.0274*<br>(0.0143) |
| % of Emergency Admissions | −0.0036<br>(0.0031) | −0.0046<br>(0.0032) | −0.0041<br>(0.0032) | −0.0051<br>(0.0032) | −0.0022<br>(0.0032) | −0.0023<br>(0.0031) |
| % of Patients Aged 60–74 | −0.0285*<br>(0.0159) | −0.0265<br>(0.0164) | −0.0352**<br>(0.0178) | −0.0362**<br>(0.0178) | −0.0416**<br>(0.0175) | −0.0400**<br>(0.0175) |
| % of Patients Aged 75+ | 0.0211***<br>(0.0070) | 0.0183**<br>(0.0072) | 0.0167**<br>(0.0073) | 0.0168**<br>(0.0072) | 0.0246***<br>(0.0074) | 0.0241***<br>(0.0073) |
| Hospital Beds 200–399 | 0.0493<br>(0.2598) | −0.0250<br>(0.2754) | −0.0419<br>(0.2756) | −0.0312<br>(0.2770) | −0.2524<br>(0.3103) | −0.2501<br>(0.3099) |
| Hospital Beds 400–599 | 0.1333<br>(0.2768) | 0.0970<br>(0.2962) | 0.2242<br>(0.2944) | 0.2337<br>(0.2953) | −0.1657<br>(0.3304) | −0.1570<br>(0.3307) |
| Hospital Beds 600–799 | −0.0876<br>(0.2823) | −0.1293<br>(0.3006) | 0.0075<br>(0.2984) | −0.0483<br>(0.3014) | −0.4670<br>(0.3358) | −0.4470<br>(0.3359) |
| Hospital Beds 800–999 | −0.1812<br>(0.2869) | −0.2327<br>(0.3043) | −0.0663<br>(0.3019) | −0.1355<br>(0.3040) | −0.5425<br>(0.3378) | −0.5363<br>(0.3379) |
| Hospital Beds 1000–1499 | −0.2095<br>(0.2978) | −0.2786<br>(0.3150) | −0.0885<br>(0.3150) | −0.1671<br>(0.3173) | −0.5614<br>(0.3498) | −0.5472<br>(0.3503) |
| Hospital Beds 1500+ | −0.6520*<br>(0.3916) | −0.6937*<br>(0.4013) | −0.5331<br>(0.4077) | −0.5424<br>(0.4073) | −0.9627**<br>(0.4336) | −0.9728**<br>(0.4355) |
| % Readmissions (16–74) |  | −0.0164<br>(0.0196) | −0.0138<br>(0.0191) | −0.0155<br>(0.0193) | −0.0062<br>(0.0230) | −0.0060<br>(0.0232) |
| % Readmissions (75+) |  | 0.0153<br>(0.0172) | 0.0152<br>(0.0169) | 0.0166<br>(0.0170) | −0.0071<br>(0.0194) | −0.0079<br>(0.0193) |
| Log Care Home Beds |  |  | −0.2542***<br>(0.0864) | −0.2053**<br>(0.0882) | −0.2910***<br>(0.0882) | −0.2718***<br>(0.0881) |
| Log Care Home Price |  |  | 0.3333<br>(0.4060) | 0.4361<br>(0.4045) | 0.4474<br>(0.4101) | 0.5479<br>(0.4130) |
| Special Staff |  |  |  | 0.0044<br>(0.0175) | −0.0068<br>(0.0171) | −0.0088<br>(0.0172) |
| Nurse Staff Satisfaction |  |  |  |  | 0.0011<br>(0.0021) | 0.0017<br>(0.0021) |
| Medical Staff Satisfaction |  |  |  |  | −0.0037**<br>(0.0015) | −0.0036**<br>(0.0015) |
| Management |  |  |  |  |  | −0.0081*<br>(0.0049) |
| 2012/13 | 0.0121<br>(0.0430) | 0.0190<br>(0.0434) | 0.0329<br>(0.0439) | 0.0340<br>(0.0434) | 0.0666<br>(0.0429) | 0.0689<br>(0.0427) |
| 2013/14 | 0.0910**<br>(0.0433) | 0.1066**<br>(0.0458) | 0.1160**<br>(0.0461) | 0.1265***<br>(0.0484) | 0.1200**<br>(0.0469) | 0.1396***<br>(0.0484) |
| 2014/15 | 0.1981***<br>(0.0453) | 0.2079***<br>(0.0460) | 0.2258***<br>(0.0468) | 0.2460***<br>(0.0486) | 0.2644***<br>(0.0470) | 0.2883***<br>(0.0491) |
| Constant | −5.6966***<br>(0.7420) | −5.3258***<br>(0.7827) | −5.3709*<br>(3.0305) | −6.2641**<br>(3.0244) | −5.1754*<br>(3.0446) | −5.7886*<br>(3.0487) |
| Observations | 812 | 792 | 774 | 773 | 753 | 752 |
| AIC | 10214.958 | 9928.507 | 9742.087 | 9715.113 | 9400.264 | 9383.748 |
| BIC | 10299.549 | 10021.998 | 9844.422 | 9822.069 | 9515.866 | 9503.940 |

\* $p < 0.10$ , \*\* $p < 0.05$ , \*\*\* $p < 0.01$ . Model 1 consists of hospital and patient level variables. Model 2 adds hospital quality variables. Model 3 adds long term care variables. Model 4 and 5 adds special staff and staff satisfaction variables respectively. Model 6 add hospitals' management quality variable. AIC: Akaike Information Criterion. BIC: Bayesian Information Criterion. Results are reported using observed information matrix standard errors (OIM) and are given in parentheses. Exposure term, log of hospital beds has a coefficient of 1.

**Table 9.4:** NHS Days of Delays

|  | *Model 1* | *Model 2* | *Model 3* | *Model 4* | *Model 5* | *Model 6* |
| --- | --- | --- | --- | --- | --- | --- |
| Teaching Trust | −0.1047<br>(0.2060) | −0.1451<br>(0.2048) | 0.0803<br>(0.2162) | 0.0211<br>(0.2211) | 0.0823<br>(0.2224) | 0.0803<br>(0.2243) |
| Foundation Trust | −0.3654***<br>(0.1110) | −0.4395***<br>(0.1140) | −0.4781***<br>(0.1164) | −0.5047***<br>(0.1173) | −0.5519***<br>(0.1158) | −0.5210***<br>(0.1172) |
| Specialist Trust | 0.5733<br>(0.4109) | 0.4212<br>(0.4381) | 0.6616<br>(0.4087) | 0.6063<br>(0.4129) | −0.0270<br>(0.4822) | −0.0248<br>(0.4817) |
| Mental Trust | −0.0524<br>(0.2509) | 0.1716<br>(0.2807) | 0.1386<br>(0.2828) | 0.0697<br>(0.2838) | −0.4528<br>(0.3331) | −0.4532<br>(0.3327) |
| % of Male Patients | 0.0124<br>(0.0155) | 0.0127<br>(0.0161) | 0.0187<br>(0.0167) | 0.0200<br>(0.0165) | 0.0298*<br>(0.0167) | 0.0302*<br>(0.0167) |
| % of Emergency Admissions | −0.0003<br>(0.0037) | −0.0006<br>(0.0039) | −0.0025<br>(0.0039) | −0.0034<br>(0.0040) | −0.0007<br>(0.0040) | −0.0011<br>(0.0040) |
| % of Patients Aged 60–74 | −0.0155<br>(0.0175) | −0.0219<br>(0.0179) | −0.0380**<br>(0.0192) | −0.0396**<br>(0.0194) | −0.0396**<br>(0.0193) | −0.0396**<br>(0.0194) |
| % of Patients Aged 75+ | 0.0215***<br>(0.0073) | 0.0233***<br>(0.0075) | 0.0213***<br>(0.0075) | 0.0217***<br>(0.0075) | 0.0272***<br>(0.0078) | 0.0273***<br>(0.0078) |
| Hospital Beds 200–399 | 0.7812***<br>(0.3018) | 0.7326**<br>(0.3389) | 0.8500***<br>(0.3229) | 0.8833***<br>(0.3284) | 0.2474<br>(0.4090) | 0.2404<br>(0.4071) |
| Hospital Beds 400–599 | 0.6377**<br>(0.3138) | 0.5659<br>(0.3498) | 0.7973**<br>(0.3338) | 0.8410**<br>(0.3401) | −0.0285<br>(0.4227) | −0.0266<br>(0.4210) |
| Hospital Beds 600–799 | 0.5957*<br>(0.3213) | 0.5334<br>(0.3571) | 0.7748**<br>(0.3408) | 0.7674**<br>(0.3494) | −0.1259<br>(0.4293) | −0.1141<br>(0.4274) |
| Hospital Beds 800–999 | 0.4151<br>(0.3266) | 0.3445<br>(0.3614) | 0.5947*<br>(0.3449) | 0.5746<br>(0.3515) | −0.3007<br>(0.4311) | −0.3069<br>(0.4296) |
| Hospital Beds 1000–1499 | 0.3277<br>(0.3359) | 0.2460<br>(0.3707) | 0.5369<br>(0.3568) | 0.5057<br>(0.3627) | −0.3537<br>(0.4407) | −0.3438<br>(0.4396) |
| Hospital Beds 1500+ | −0.3950<br>(0.4391) | −0.3910<br>(0.4598) | −0.1839<br>(0.4579) | −0.1535<br>(0.4626) | −1.0332*<br>(0.5302) | −1.0660**<br>(0.5314) |
| % Readmissions (16–74) |  | −0.0535**<br>(0.0258) | −0.0638**<br>(0.0251) | −0.0654***<br>(0.0251) | −0.0348<br>(0.0304) | −0.0356<br>(0.0307) |
| % of Readmissions (75+) |  | 0.0508**<br>(0.0216) | 0.0633***<br>(0.0212) | 0.0653***<br>(0.0212) | 0.0279<br>(0.0251) | 0.0285<br>(0.0250) |
| Log Care Home Beds |  |  | −0.3243***<br>(0.0873) | −0.2955***<br>(0.0882) | −0.3676***<br>(0.0889) | −0.3433***<br>(0.0893) |
| Log Care Home Price |  |  | −0.2012<br>(0.3887) | −0.1737<br>(0.3875) | −0.0454<br>(0.3928) | 0.0827<br>(0.3961) |
| Special Staff |  |  |  | −0.0062<br>(0.0198) | −0.0186<br>(0.0194) | −0.0202<br>(0.0194) |
| Nurse Staff Satisfaction |  |  |  |  | −0.0008<br>(0.0023) | −0.0000<br>(0.0023) |
| Medical Staff Satisfaction |  |  |  |  | −0.0044***<br>(0.0017) | −0.0043**<br>(0.0017) |
| Management |  |  |  |  |  | −0.0110*<br>(0.0057) |
| 2012/13 | 0.0588<br>(0.0515) | 0.0534<br>(0.0520) | 0.0614<br>(0.0520) | 0.0624<br>(0.0517) | 0.1048**<br>(0.0509) | 0.1064**<br>(0.0505) |
| 2013/14 | 0.1901***<br>(0.0513) | 0.1930***<br>(0.0537) | 0.1922***<br>(0.0537) | 0.1921***<br>(0.0565) | 0.1763***<br>(0.0549) | 0.2026***<br>(0.0565) |
| 2014/15 | 0.3200***<br>(0.0526) | 0.3186***<br>(0.0533) | 0.3308***<br>(0.0534) | 0.3428***<br>(0.0555) | 0.3494***<br>(0.0541) | 0.3841***<br>(0.0568) |
| Constant | −6.2792***<br>(0.8068) | −6.2914***<br>(0.8727) | −2.6123<br>(2.9801) | −2.9580<br>(2.9727) | −2.3489<br>(3.0135) | −3.1036<br>(3.0142) |
| Observations | 804 | 784 | 766 | 765 | 745 | 744 |
| AIC | 9650.276 | 9380.215 | 9205.297 | 9185.073 | 8900.940 | 8885.576 |
| BIC | 9734.689 | 9473.504 | 9307.403 | 9291.790 | 9016.275 | 9005.489 |

\* $p < 0.10$ , \*\* $p < 0.05$ , \*\*\* $p < 0.01$ . Model 1 consists of hospital and patient level variables. Model 2 adds hospital quality variables. Model 3 adds long term care variables. Model 4 and 5 adds special staff and staff satisfaction variables respectively. Model 6 add hospitals' management quality variable. AIC: Akaike Information Criterion. BIC: Bayesian Information Criterion. Results are reported using observed information matrix standard errors (OIM) and are given in parentheses. Exposure term, log of hospital beds has a coefficient of 1.

**Table 9.5:**
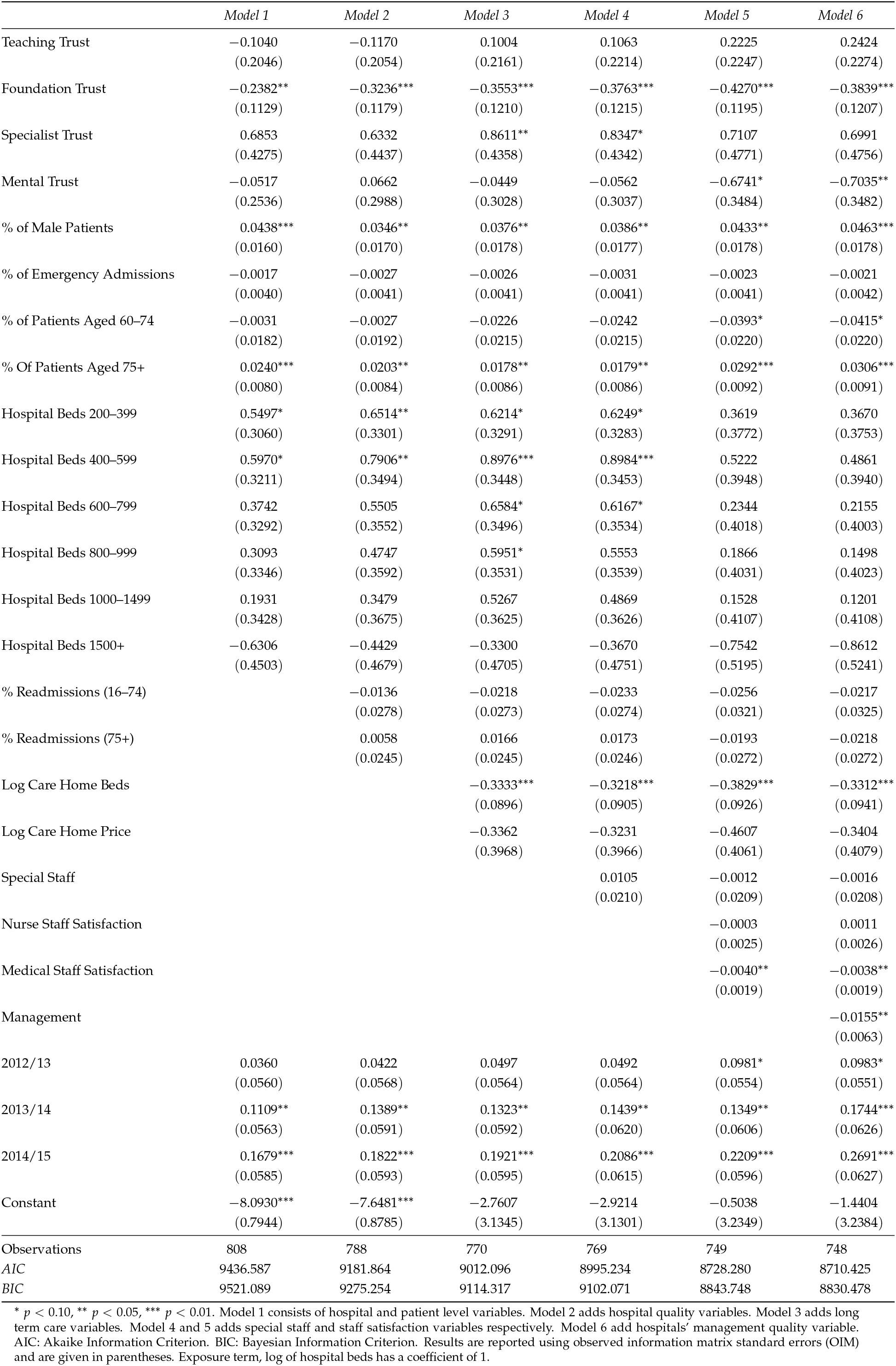
Days of Delays by Major Causes

|  | *Model 1* | *Model 2* | *Model 3* | *Model 4* | *Model 5* | *Model 6* |
| --- | --- | --- | --- | --- | --- | --- |
| Teaching Trust | −0.1040<br>(0.2046) | −0.1170<br>(0.2054) | 0.1004<br>(0.2161) | 0.1063<br>(0.2214) | 0.2225<br>(0.2247) | 0.2424<br>(0.2274) |
| Foundation Trust | −0.2382**<br>(0.1129) | −0.3236***<br>(0.1179) | −0.3553***<br>(0.1210) | −0.3763***<br>(0.1215) | −0.4270***<br>(0.1195) | −0.3839***<br>(0.1207) |
| Specialist Trust | 0.6853<br>(0.4275) | 0.6332<br>(0.4437) | 0.8611**<br>(0.4358) | 0.8347*<br>(0.4342) | 0.7107<br>(0.4771) | 0.6991<br>(0.4756) |
| Mental Trust | −0.0517<br>(0.2536) | 0.0662<br>(0.2988) | −0.0449<br>(0.3028) | −0.0562<br>(0.3037) | −0.6741*<br>(0.3484) | −0.7035**<br>(0.3482) |
| % of Male Patients | 0.0438***<br>(0.0160) | 0.0346**<br>(0.0170) | 0.0376**<br>(0.0178) | 0.0386**<br>(0.0177) | 0.0433**<br>(0.0178) | 0.0463***<br>(0.0178) |
| % of Emergency Admissions | −0.0017<br>(0.0040) | −0.0027<br>(0.0041) | −0.0026<br>(0.0041) | −0.0031<br>(0.0041) | −0.0023<br>(0.0041) | −0.0021<br>(0.0042) |
| % of Patients Aged 60–74 | −0.0031<br>(0.0182) | −0.0027<br>(0.0192) | −0.0226<br>(0.0215) | −0.0242<br>(0.0215) | −0.0393*<br>(0.0220) | −0.0415*<br>(0.0220) |
| % Of Patients Aged 75+ | 0.0240***<br>(0.0080) | 0.0203**<br>(0.0084) | 0.0178**<br>(0.0086) | 0.0179**<br>(0.0086) | 0.0292***<br>(0.0092) | 0.0306***<br>(0.0091) |
| Hospital Beds 200–399 | 0.5497*<br>(0.3060) | 0.6514**<br>(0.3301) | 0.6214*<br>(0.3291) | 0.6249*<br>(0.3283) | 0.3619<br>(0.3772) | 0.3670<br>(0.3753) |
| Hospital Beds 400–599 | 0.5970*<br>(0.3211) | 0.7906**<br>(0.3494) | 0.8976***<br>(0.3448) | 0.8984***<br>(0.3453) | 0.5222<br>(0.3948) | 0.4861<br>(0.3940) |
| Hospital Beds 600–799 | 0.3742<br>(0.3292) | 0.5505<br>(0.3552) | 0.6584*<br>(0.3496) | 0.6167*<br>(0.3534) | 0.2344<br>(0.4018) | 0.2155<br>(0.4003) |
| Hospital Beds 800–999 | 0.3093<br>(0.3346) | 0.4747<br>(0.3592) | 0.5951*<br>(0.3531) | 0.5553<br>(0.3539) | 0.1866<br>(0.4031) | 0.1498<br>(0.4023) |
| Hospital Beds 1000–1499 | 0.1931<br>(0.3428) | 0.3479<br>(0.3675) | 0.5267<br>(0.3625) | 0.4869<br>(0.3626) | 0.1528<br>(0.4107) | 0.1201<br>(0.4108) |
| Hospital Beds 1500+ | −0.6306<br>(0.4503) | −0.4429<br>(0.4679) | −0.3300<br>(0.4705) | −0.3670<br>(0.4751) | −0.7542<br>(0.5195) | −0.8612<br>(0.5241) |
| % Readmissions (16–74) |  | −0.0136<br>(0.0278) | −0.0218<br>(0.0273) | −0.0233<br>(0.0274) | −0.0256<br>(0.0321) | −0.0217<br>(0.0325) |
| % Readmissions (75+) |  | 0.0058<br>(0.0245) | 0.0166<br>(0.0245) | 0.0173<br>(0.0246) | −0.0193<br>(0.0272) | −0.0218<br>(0.0272) |
| Log Care Home Beds |  |  | −0.3333***<br>(0.0896) | −0.3218***<br>(0.0905) | −0.3829***<br>(0.0926) | −0.3312***<br>(0.0941) |
| Log Care Home Price |  |  | −0.3362<br>(0.3968) | −0.3231<br>(0.3966) | −0.4607<br>(0.4061) | −0.3404<br>(0.4079) |
| Special Staff |  |  |  | 0.0105<br>(0.0210) | −0.0012<br>(0.0209) | −0.0016<br>(0.0208) |
| Nurse Staff Satisfaction |  |  |  |  | −0.0003<br>(0.0025) | 0.0011<br>(0.0026) |
| Medical Staff Satisfaction |  |  |  |  | −0.0040**<br>(0.0019) | −0.0038**<br>(0.0019) |
| Management |  |  |  |  |  | −0.0155**<br>(0.0063) |
| 2012/13 | 0.0360<br>(0.0560) | 0.0422<br>(0.0568) | 0.0497<br>(0.0564) | 0.0492<br>(0.0564) | 0.0981*<br>(0.0554) | 0.0983*<br>(0.0551) |
| 2013/14 | 0.1109**<br>(0.0563) | 0.1389**<br>(0.0591) | 0.1323**<br>(0.0592) | 0.1439**<br>(0.0620) | 0.1349**<br>(0.0606) | 0.1744***<br>(0.0626) |
| 2014/15 | 0.1679***<br>(0.0585) | 0.1822***<br>(0.0593) | 0.1921***<br>(0.0595) | 0.2086***<br>(0.0615) | 0.2209***<br>(0.0596) | 0.2691***<br>(0.0627) |
| Constant | −8.0930***<br>(0.7944) | −7.6481***<br>(0.8785) | −2.7607<br>(3.1345) | −2.9214<br>(3.1301) | −0.5038<br>(3.2349) | −1.4404<br>(3.2384) |
| Observations | 808 | 788 | 770 | 769 | 749 | 748 |
| AIC | 9436.587 | 9181.864 | 9012.096 | 8995.234 | 8728.280 | 8710.425 |
| BIC | 9521.089 | 9275.254 | 9114.317 | 9102.071 | 8843.748 | 8830.478 |

\* $p < 0.10$ , \*\* $p < 0.05$ , \*\*\* $p < 0.01$ . Model 1 consists of hospital and patient level variables. Model 2 adds hospital quality variables. Model 3 adds long term care variables. Model 4 and 5 adds special staff and staff satisfaction variables respectively. Model 6 add hospitals' management quality variable. AIC: Akaike Information Criterion. BIC: Bayesian Information Criterion. Results are reported using observed information matrix standard errors (OIM) and are given in parentheses. Exposure term, log of hospital beds has a coefficient of 1.

**Table 9.6:** Days of Delay For Major Causes (NHS)

|  | *Model 1* | *Model 2* | *Model 3* | *Model 4* | *Model 5* | *Model 6* |
| --- | --- | --- | --- | --- | --- | --- |
| Teaching Trust | 0.0425<br>(0.1980) | 0.0400<br>(0.1992) | 0.2530<br>(0.2082) | 0.1895<br>(0.2142) | 0.2530<br>(0.2163) | 0.2492<br>(0.2172) |
| Foundation Trust | -0.4755***<br>(0.1179) | -0.5148***<br>(0.1228) | -0.5622***<br>(0.1238) | -0.5638***<br>(0.1247) | -0.5855***<br>(0.1242) | -0.5506***<br>(0.1263) |
| Specialist Trust | 1.4628***<br>(0.4165) | 1.4433***<br>(0.4261) | 1.6662***<br>(0.4156) | 1.6407***<br>(0.4182) | 1.5197***<br>(0.4337) | 1.5381***<br>(0.4334) |
| Mental Trust | -0.3003<br>(0.2726) | -0.2297<br>(0.3195) | -0.4998<br>(0.3267) | -0.5484*<br>(0.3274) | -1.1760***<br>(0.3829) | -1.2135***<br>(0.3829) |
| % of Male Patients | 0.0248<br>(0.0177) | 0.0250<br>(0.0186) | 0.0359*<br>(0.0193) | 0.0364*<br>(0.0192) | 0.0394**<br>(0.0192) | 0.0423**<br>(0.0193) |
| % of Emergency Admissions | -0.0008<br>(0.0043) | 0.0001<br>(0.0045) | -0.0004<br>(0.0046) | -0.0011<br>(0.0046) | 0.0003<br>(0.0046) | 0.0005<br>(0.0047) |
| % of Patients Aged 60–74 | -0.0146<br>(0.0191) | -0.0161<br>(0.0201) | -0.0535**<br>(0.0223) | -0.0514**<br>(0.0225) | -0.0585**<br>(0.0228) | -0.0616***<br>(0.0230) |
| % of Patients Aged 75+ | 0.0266***<br>(0.0081) | 0.0267***<br>(0.0084) | 0.0260***<br>(0.0086) | 0.0261***<br>(0.0086) | 0.0336***<br>(0.0091) | 0.0353***<br>(0.0091) |
| Hospital Beds 200–399 | 0.3713<br>(0.2918) | 0.4365<br>(0.3109) | 0.6640**<br>(0.3123) | 0.7016**<br>(0.3186) | 0.5873*<br>(0.3444) | 0.5903*<br>(0.3439) |
| Hospital Beds 400–599 | 0.2937<br>(0.3074) | 0.3293<br>(0.3268) | 0.6594**<br>(0.3285) | 0.7134**<br>(0.3351) | 0.4591<br>(0.3645) | 0.4410<br>(0.3641) |
| Hospital Beds 600–799 | 0.2565<br>(0.3166) | 0.2934<br>(0.3346) | 0.6092*<br>(0.3338) | 0.6533*<br>(0.3430) | 0.3882<br>(0.3713) | 0.3747<br>(0.3705) |
| Hospital Beds 800–999 | 0.1725<br>(0.3225) | 0.2068<br>(0.3402) | 0.4957<br>(0.3375) | 0.5218<br>(0.3446) | 0.2695<br>(0.3726) | 0.2419<br>(0.3725) |
| Hospital Beds 1000–1499 | 0.0840<br>(0.3320) | 0.1175<br>(0.3506) | 0.4276<br>(0.3484) | 0.4395<br>(0.3542) | 0.2217<br>(0.3823) | 0.2062<br>(0.3824) |
| Hospital Beds 1500+ | -0.5417<br>(0.4394) | -0.4709<br>(0.4529) | -0.2387<br>(0.4546) | -0.1769<br>(0.4596) | -0.4510<br>(0.4843) | -0.5024<br>(0.4859) |
| % Readmissions (16–74) |  | -0.0258<br>(0.0304) | -0.0434<br>(0.0298) | -0.0443<br>(0.0297) | -0.0353<br>(0.0350) | -0.0337<br>(0.0352) |
| % Readmissions (75+) |  | 0.0192<br>(0.0261) | 0.0396<br>(0.0264) | 0.0419<br>(0.0263) | -0.0020<br>(0.0294) | -0.0023<br>(0.0294) |
| Log Care Home Beds |  |  | -0.3534***<br>(0.0920) | -0.3342***<br>(0.0927) | -0.3554***<br>(0.0921) | -0.3139***<br>(0.0945) |
| Log Care Home Price |  |  | -1.1780***<br>(0.4001) | -1.1923***<br>(0.3981) | -1.1960***<br>(0.4058) | -1.0974***<br>(0.4084) |
| Special Staff |  |  |  | -0.0241<br>(0.0231) | -0.0347<br>(0.0230) | -0.0354<br>(0.0229) |
| Nurse Staff Satisfaction |  |  |  |  | 0.0007<br>(0.0027) | 0.0017<br>(0.0028) |
| Medical Staff Satisfaction |  |  |  |  | -0.0049**<br>(0.0020) | -0.0048**<br>(0.0020) |
| Management |  |  |  |  |  | -0.0117*<br>(0.0069) |
| 2012/13 | 0.1244*<br>(0.0635) | 0.1189*<br>(0.0645) | 0.1172*<br>(0.0639) | 0.1170*<br>(0.0639) | 0.1631***<br>(0.0632) | 0.1628***<br>(0.0630) |
| 2013/14 | 0.2494***<br>(0.0636) | 0.2561***<br>(0.0672) | 0.2462***<br>(0.0666) | 0.2256***<br>(0.0698) | 0.2116***<br>(0.0689) | 0.2409***<br>(0.0713) |
| 2014/15 | 0.3035***<br>(0.0652) | 0.3051***<br>(0.0666) | 0.3122***<br>(0.0661) | 0.3015***<br>(0.0685) | 0.3134***<br>(0.0673) | 0.3484***<br>(0.0707) |
| Constant | -7.1328***<br>(0.8302) | -7.1943***<br>(0.9205) | 2.9665<br>(3.1344) | 2.9176<br>(3.1235) | 4.0081<br>(3.1558) | 3.2104<br>(3.1700) |
| Observations | 800 | 780 | 762 | 761 | 741 | 740 |
| AIC | 9096.669 | 8857.894 | 8689.685 | 8673.441 | 8426.435 | 8412.406 |
| BIC | 9180.992 | 8951.080 | 8791.676 | 8780.037 | 8541.635 | 8532.179 |

\* $p < 0.10$ , \*\* $p < 0.05$ , \*\*\* $p < 0.01$ . Model 1 consists of hospital and patient level variables. Model 2 adds hospital quality variables. Model 3 adds long term care variables. Model 4 and 5 adds special staff and staff satisfaction variables respectively. Model 6 add hospitals' management quality variable. AIC: Akaike Information Criterion. BIC: Bayesian Information Criterion. Results are reported using observed information matrix standard errors (OIM) and are given in parentheses. Exposure term, log of hospital beds has a coefficient of 1.

**Table 9.7:** Days of Delays & Staff Satisfaction Results

|  | (1)<br>*Total* | (2)<br>*NHS* | (3)<br>*Major* | (4)<br>*Major (NHS)* |
| --- | --- | --- | --- | --- |
| Teaching Trust | 0.0147<br>(0.1414) | 0.1097<br>(0.1489) | −0.0109<br>(0.1592) | 0.2430<br>(0.2176) |
| Foundation Trust | −0.2503***<br>(0.0756) | −0.3788***<br>(0.0843) | −0.2186**<br>(0.0889) | −0.5472***<br>(0.1262) |
| Specialist Trust | 0.2267<br>(0.2972) | 0.0305<br>(0.3368) | 0.4017<br>(0.3629) | 1.5552***<br>(0.4334) |
| Mental Trust | −0.0006<br>(0.2201) | −0.5949**<br>(0.2772) | −1.0061***<br>(0.3029) | −1.2203***<br>(0.3835) |
| % of Male Patients | 0.0126<br>(0.0115) | 0.0234*<br>(0.0133) | 0.0473***<br>(0.0141) | 0.0425**<br>(0.0193) |
| % of Emergency Admissions | −0.0018<br>(0.0025) | −0.0006<br>(0.0033) | 0.0003<br>(0.0034) | 0.0003<br>(0.0046) |
| % of Patients Aged 60–74 | −0.0437***<br>(0.0120) | −0.0513***<br>(0.0147) | −0.0494***<br>(0.0162) | −0.0607***<br>(0.0230) |
| % of Patients Aged 75+ | 0.0213***<br>(0.0050) | 0.0240***<br>(0.0057) | 0.0185***<br>(0.0065) | 0.0349***<br>(0.0091) |
| Hospital Beds 200–399 | −0.2378<br>(0.2134) | 0.4266<br>(0.2660) | 0.3738<br>(0.2725) | 0.5979*<br>(0.3449) |
| Hospital Beds 400–599 | −0.1088<br>(0.2236) | 0.2270<br>(0.2734) | 0.4850*<br>(0.2868) | 0.4450<br>(0.3654) |
| Hospital Beds 600–799 | −0.1992<br>(0.2281) | 0.3504<br>(0.2794) | 0.4115<br>(0.2912) | 0.3673<br>(0.3714) |
| Hospital Beds 800–1000 | −0.2012<br>(0.2319) | 0.2130<br>(0.2831) | 0.4844*<br>(0.2942) | 0.2360<br>(0.3735) |
| Hospital Beds 1000–1499 | −0.1777<br>(0.2401) | 0.2412<br>(0.2913) | 0.4599<br>(0.3033) | 0.2065<br>(0.3834) |
| Hospital Beds 1500+ | −0.5722*<br>(0.2966) | −0.2858<br>(0.3522) | −0.0564<br>(0.3647) | −0.5001<br>(0.4879) |
| % Readmissions (16–74) | 0.0232<br>(0.0199) | −0.0018<br>(0.0258) | 0.0160<br>(0.0284) | −0.0340<br>(0.0353) |
| % Readmissions (75+) | −0.0333*<br>(0.0173) | −0.0039<br>(0.0219) | −0.0338<br>(0.0240) | −0.0023<br>(0.0294) |
| Log Care Home Beds | −0.2925***<br>(0.0533) | −0.3171***<br>(0.0597) | −0.2709***<br>(0.0662) | −0.3097***<br>(0.0946) |
| Log Care Home Price | 0.5140**<br>(0.2528) | −0.0605<br>(0.2715) | −0.4970*<br>(0.2820) | −1.0674***<br>(0.4060) |
| Special Staff | −0.0237<br>(0.0158) | −0.0436**<br>(0.0178) | −0.0199<br>(0.0190) | −0.0359<br>(0.0229) |
| Medical Staff Satisfaction | −0.0046***<br>(0.0014) | −0.0052***<br>(0.0016) | −0.0046**<br>(0.0018) | −0.0046**<br>(0.0020) |
| Management | −0.0121***<br>(0.0045) | −0.0121**<br>(0.0052) | −0.0123**<br>(0.0058) | −0.0108<br>(0.0068) |
| 2012/13 | 0.0864**<br>(0.0427) | 0.1109**<br>(0.0506) | 0.0885<br>(0.0559) | 0.1688***<br>(0.0623) |
| 2013/14 | 0.1432***<br>(0.0471) | 0.1875***<br>(0.0548) | 0.1334**<br>(0.0619) | 0.2400***<br>(0.0713) |
| 2014/15 | 0.3012***<br>(0.0459) | 0.3653***<br>(0.0541) | 0.2453***<br>(0.0612) | 0.3460***<br>(0.0705) |
| Constant | −4.6836**<br>(1.9004) | −2.0327<br>(2.0727) | −6.7853***<br>(2.2215) | 3.0358<br>(3.1674) |
| Observations | 753 | 753 | 753 | 740 |

\* $p < 0.10$ , \*\* $p < 0.05$ , \*\*\* $p < 0.01$ . Results are reported using observed information matrix standard errors (OIM) and are given in parentheses. Exposure term, log of hospital beds has a coefficient of 1 in models (1), (2) and (4). Due to convergence issues, model (3) for delayed days due to major causes uses log of hospital bed–days as the exposure term.

**Table 9.8:** Days of Delays & Staff Satisfaction Results (Without Specialists)

|  | (1)<br>*Total* | (2)<br>*NHS* | (3)<br>*Major* | (4)<br>*Major (NHS)* |
| --- | --- | --- | --- | --- |
| Teaching Trust | 0.0625<br>(0.1388) | 0.0402<br>(0.1505) | 0.1321<br>(0.1616) | 0.1227<br>(0.1634) |
| Foundation Trust | -0.2520***<br>(0.0751) | -0.4077***<br>(0.0851) | -0.2993***<br>(0.0916) | -0.3355***<br>(0.0960) |
| Mental Trust | -0.0309<br>(0.2247) | -0.5645*<br>(0.2916) | -0.9786***<br>(0.3120) | -1.2596***<br>(0.3489) |
| % of Male Patients | 0.0221*<br>(0.0124) | 0.0071<br>(0.0144) | 0.0268*<br>(0.0162) | 0.0313*<br>(0.0175) |
| % of Emergency Admissions | -0.0012<br>(0.0026) | 0.0027<br>(0.0033) | -0.0044<br>(0.0035) | 0.0045<br>(0.0038) |
| % of Patients Aged 60–74 | -0.0418***<br>(0.0138) | -0.0384**<br>(0.0162) | -0.0598***<br>(0.0182) | -0.0688***<br>(0.0193) |
| % of Patients Aged 75+ | 0.0222***<br>(0.0052) | 0.0046<br>(0.0060) | 0.0268***<br>(0.0068) | 0.0129*<br>(0.0073) |
| Hospital Beds 200–399 | 0.2017<br>(0.3184) | 0.5323<br>(0.3514) | 0.4778<br>(0.3849) | 1.0127**<br>(0.4174) |
| Hospital Beds 400–599 | 0.3438<br>(0.3211) | 0.4288<br>(0.3502) | 0.5540<br>(0.3870) | 0.8742**<br>(0.4169) |
| Hospital Beds 600–799 | 0.2569<br>(0.3238) | 0.5571<br>(0.3562) | 0.4617<br>(0.3885) | 0.9844**<br>(0.4208) |
| Hospital Beds 800–999 | 0.2667<br>(0.3265) | 0.4501<br>(0.3597) | 0.5175<br>(0.3910) | 0.9836**<br>(0.4235) |
| Hospital Beds 1000–1499 | 0.2965<br>(0.3323) | 0.4982<br>(0.3658) | 0.4950<br>(0.3973) | 1.0136**<br>(0.4310) |
| Hospital Beds 1500+ | -0.0760<br>(0.3773) | 0.0539<br>(0.4166) | -0.1676<br>(0.4531) | 0.6624<br>(0.4786) |
| % Readmissions (16–74) | 0.0168<br>(0.0207) | 0.0235<br>(0.0272) | 0.0070<br>(0.0297) | 0.0058<br>(0.0336) |
| % Readmissions (75+) | -0.0293<br>(0.0181) | -0.0181<br>(0.0235) | -0.0472*<br>(0.0256) | -0.0217<br>(0.0285) |
| Log Care Home Beds | -0.3643***<br>(0.0597) | -0.3676***<br>(0.0655) | -0.2464***<br>(0.0727) | -0.3237***<br>(0.0777) |
| Log Care Home Price | 0.5635**<br>(0.2518) | -0.0119<br>(0.2748) | -0.4858*<br>(0.2953) | -1.1968***<br>(0.3111) |
| Special Staff | -0.0282*<br>(0.0160) | -0.0319*<br>(0.0187) | -0.0233<br>(0.0200) | -0.0404*<br>(0.0229) |
| Medical Staff Satisfaction | -0.0046***<br>(0.0014) | -0.0052***<br>(0.0016) | -0.0041**<br>(0.0018) | -0.0045**<br>(0.0020) |
| Management | -0.0115**<br>(0.0045) | -0.0122**<br>(0.0052) | -0.0147**<br>(0.0059) | -0.0065<br>(0.0065) |
| 2012/13 | 0.0864**<br>(0.0433) | 0.1107**<br>(0.0511) | 0.1134**<br>(0.0569) | 0.1452**<br>(0.0664) |
| 2013/14 | 0.1379***<br>(0.0477) | 0.1995***<br>(0.0559) | 0.1795***<br>(0.0634) | 0.2189***<br>(0.0729) |
| 2014/15 | 0.2955***<br>(0.0464) | 0.3718***<br>(0.0552) | 0.2874***<br>(0.0627) | 0.3243***<br>(0.0720) |
| Constant | -5.4282***<br>(1.9543) | -7.2813***<br>(2.1674) | 0.1535<br>(2.4408) | -1.9061<br>(2.5555) |
| Observations | 713 | 713 | 713 | 713 |

\* $p < 0.10$ , \*\* $p < 0.05$ , \*\*\* $p < 0.01$ . Results are reported using observed information matrix standard errors (OIM). Exposure term, log of hospital beds has a coefficient of 1 in models (1) and (3). Due to convergence issues, models (2) and (4) for NHS related delayed days used log of hospital bed-days as the exposure term.

**Table 9.9:** Correlated Random Effects & Staff Satisfaction Results

|  | (1)<br>*Total* | (2)<br>*NHS* | (3)<br>*Major* | (4)<br>*Major (NHS)* |
| --- | --- | --- | --- | --- |
| Teaching Trust | 0.0277<br>(0.1412) | 0.1394<br>(0.1504) | 0.1029<br>(0.1600) | 0.0682<br>(0.1615) |
| Foundation Trust | -0.2099***<br>(0.0808) | -0.3277***<br>(0.0907) | -0.2198**<br>(0.0956) | -0.2714***<br>(0.0996) |
| Specialist Trust | 0.3470<br>(0.3096) | 0.0267<br>(0.3448) | 0.3274<br>(0.3751) | 0.7039*<br>(0.3711) |
| Mental Trust | 0.0202<br>(0.2196) | -0.6136**<br>(0.2751) | -1.0158***<br>(0.2975) | -1.1413***<br>(0.3029) |
| % of Male Patients | 0.0143<br>(0.0114) | 0.0225*<br>(0.0134) | 0.0435***<br>(0.0147) | 0.0413***<br>(0.0154) |
| % of Emergency Admissions | -0.0019<br>(0.0026) | 0.0001<br>(0.0034) | -0.0033<br>(0.0035) | 0.0062<br>(0.0038) |
| % of Patients Aged 60–74 | -0.0441***<br>(0.0122) | -0.0469***<br>(0.0149) | -0.0578***<br>(0.0165) | -0.0447***<br>(0.0172) |
| % of Patients Aged 75+ | 0.0213***<br>(0.0050) | 0.0230***<br>(0.0057) | 0.0310***<br>(0.0066) | 0.0136*<br>(0.0069) |
| Hospital Beds 200–399 | -0.2192<br>(0.2150) | 0.3609<br>(0.2702) | 0.3390<br>(0.2868) | 0.5506**<br>(0.2795) |
| Hospital Beds 400–599 | -0.0926<br>(0.2272) | 0.1469<br>(0.2792) | 0.3716<br>(0.3026) | 0.4097<br>(0.2973) |
| Hospital Beds 600–799 | -0.1959<br>(0.2311) | 0.2651<br>(0.2842) | 0.2917<br>(0.3052) | 0.5551*<br>(0.3021) |
| Hospital Beds 800–999 | -0.1953<br>(0.2354) | 0.1251<br>(0.2883) | 0.3356<br>(0.3083) | 0.5236*<br>(0.3065) |
| Hospital Beds 1000–1499 | -0.1708<br>(0.2435) | 0.1524<br>(0.2967) | 0.3132<br>(0.3161) | 0.5278*<br>(0.3160) |
| Hospital Beds 1500+ | -0.5707*<br>(0.3014) | -0.4140<br>(0.3624) | -0.3570<br>(0.3834) | 0.1416<br>(0.3809) |
| % Readmissions (16–74) | 0.0226<br>(0.0199) | 0.0044<br>(0.0262) | 0.0078<br>(0.0283) |  |
| % Readmissions (75+) | -0.0313*<br>(0.0172) | -0.0077<br>(0.0220) | -0.0387<br>(0.0241) |  |
| % Readmissions (16+) |  |  |  | -0.0088<br>(0.0172) |
| Log Care Home Beds | -0.2616***<br>(0.0568) | -0.3173***<br>(0.0636) | -0.2452***<br>(0.0697) | -0.3309***<br>(0.0716) |
| Log Care Home Price | 0.6007**<br>(0.2586) | -0.0496<br>(0.2801) | -0.5286*<br>(0.2920) | -1.2356***<br>(0.3019) |
| Special Staff | -0.0275*<br>(0.0158) | -0.0434**<br>(0.0180) | -0.0257<br>(0.0191) | -0.0514**<br>(0.0216) |
| *Within-Hospital Effects* |  |  |  |  |
| Medical Staff Satisfaction | -0.0040***<br>(0.0015) | -0.0040**<br>(0.0017) | -0.0038*<br>(0.0020) | -0.0043*<br>(0.0022) |
| Management | -0.0085*<br>(0.0050) | -0.0106*<br>(0.0058) | -0.0157**<br>(0.0068) | -0.0150*<br>(0.0077) |
| *Averages* |  |  |  |  |
| Medical Staff Satisfaction | -0.0015<br>(0.0048) | -0.0089<br>(0.0055) | -0.0055<br>(0.0057) | -0.0085<br>(0.0060) |
| Management | -0.0161<br>(0.0127) | 0.0069<br>(0.0144) | 0.0110<br>(0.0141) | 0.0385**<br>(0.0150) |
| 2012/13 | 0.0831*<br>(0.0425) | 0.1069**<br>(0.0504) | 0.0999*<br>(0.0561) | 0.1428**<br>(0.0649) |
| 2013/14 | 0.1271***<br>(0.0479) | 0.1830***<br>(0.0556) | 0.1533**<br>(0.0631) | 0.2155***<br>(0.0713) |
| 2014/15 | 0.2822***<br>(0.0469) | 0.3517***<br>(0.0555) | 0.2575***<br>(0.0630) | 0.3224***<br>(0.0719) |
| Constant | -5.1697***<br>(1.9298) | -1.9420<br>(2.1155) | -0.4725<br>(2.2972) | -2.7323<br>(2.3345) |
| Observations | 753 | 753 | 753 | 762 |

\* $p < 0.10$ , \*\* $p < 0.05$ , \*\*\* $p < 0.01$ . Results are reported using observed information matrix standard errors (OIM) and are given in parentheses. Exposure term, log of hospital beds has a coefficient of 1 in models (1), (2) and (3). Due to convergence issues, model (4) for NHS delayed days due to major causes uses log of hospital bed-days as the exposure term and a single readmission rate variable for ages 16 and above.

**Table 9.10:** Correlated Random Effects & Staff Satisfaction Results (without Specialists)

|  | (1)<br>*Total* | (2)<br>*NHS* | (3)<br>*Major* | (4)<br>*Major (NHS)* |
| --- | --- | --- | --- | --- |
| Teaching Trust | 0.0756<br>(0.1388) | 0.1632<br>(0.1512) | 0.1295<br>(0.1612) | 0.1269<br>(0.1643) |
| Foundation Trust | -0.2152***<br>(0.0804) | -0.3920***<br>(0.0921) | -0.3069***<br>(0.0976) | -0.3801***<br>(0.1020) |
| Mental Trust | -0.0319<br>(0.2236) | -0.5229*<br>(0.2835) | -0.9926***<br>(0.3121) | -1.2484***<br>(0.3488) |
| % of Male Patients | 0.0231*<br>(0.0124) | 0.0075<br>(0.0147) | 0.0254<br>(0.0164) | 0.0270<br>(0.0176) |
| % of Emergency Admissions | -0.0011<br>(0.0026) | -0.0010<br>(0.0034) | -0.0042<br>(0.0035) | 0.0047<br>(0.0038) |
| % of Patients Aged 60–74 | -0.0406***<br>(0.0141) | -0.0444***<br>(0.0164) | -0.0554***<br>(0.0189) | -0.0589***<br>(0.0201) |
| % of Patients Aged 75+ | 0.0220***<br>(0.0052) | 0.0188***<br>(0.0060) | 0.0259***<br>(0.0069) | 0.0109<br>(0.0074) |
| Hospital Beds 200–399 | 0.2007<br>(0.3183) | 0.4753<br>(0.3574) | 0.4438<br>(0.3866) | 0.9446**<br>(0.4210) |
| Hospital Beds 400–599 | 0.3323<br>(0.3216) | 0.2864<br>(0.3567) | 0.5179<br>(0.3899) | 0.8175*<br>(0.4217) |
| Hospital Beds 600–799 | 0.2391<br>(0.3241) | 0.4107<br>(0.3610) | 0.4370<br>(0.3908) | 0.9713**<br>(0.4250) |
| Hospital Beds 800–999 | 0.2492<br>(0.3272) | 0.2794<br>(0.3646) | 0.4863<br>(0.3933) | 0.9533**<br>(0.4277) |
| Hospital Beds 1000–1499 | 0.2791<br>(0.3331) | 0.3166<br>(0.3710) | 0.4654<br>(0.3995) | 0.9753**<br>(0.4356) |
| Hospital Beds 1500+ | -0.1099<br>(0.3797) | -0.2652<br>(0.4276) | -0.2144<br>(0.4581) | 0.5847<br>(0.4882) |
| % Readmissions (16–74) | 0.0182<br>(0.0208) | 0.0096<br>(0.0270) | 0.0102<br>(0.0303) | 0.0090<br>(0.0344) |
| % Readmissions (75+) | -0.0293<br>(0.0180) | -0.0167<br>(0.0231) | -0.0503*<br>(0.0260) | -0.0257<br>(0.0291) |
| Log Care Home Beds | -0.3520***<br>(0.0630) | -0.3370***<br>(0.0686) | -0.2660***<br>(0.0763) | -0.3871***<br>(0.0815) |
| Log Care Home Price | 0.5925**<br>(0.2574) | 0.0215<br>(0.2843) | -0.5177*<br>(0.2983) | -1.3290***<br>(0.3181) |
| Special Staff | -0.0301*<br>(0.0161) | -0.0378**<br>(0.0187) | -0.0229<br>(0.0200) | -0.0391*<br>(0.0229) |
| *Within-Hospital Effects* |  |  |  |  |
| Medical Staff Satisfaction | -0.0040***<br>(0.0015) | -0.0041**<br>(0.0018) | -0.0039*<br>(0.0020) | -0.0046**<br>(0.0022) |
| Management | -0.0095*<br>(0.0050) | -0.0117**<br>(0.0058) | -0.0172**<br>(0.0068) | -0.0161**<br>(0.0077) |
| *Averages* |  |  |  |  |
| Medical Staff Satisfaction | -0.0041<br>(0.0048) | -0.0065<br>(0.0057) | -0.0035<br>(0.0058) | -0.0057<br>(0.0062) |
| Management | -0.0037<br>(0.0128) | 0.0051<br>(0.0149) | 0.0142<br>(0.0145) | 0.0401***<br>(0.0154) |
| 2012/13 | 0.0839*<br>(0.0432) | 0.1182**<br>(0.0510) | 0.1150**<br>(0.0570) | 0.1498**<br>(0.0660) |
| 2013/14 | 0.1295***<br>(0.0486) | 0.2083***<br>(0.0567) | 0.1864***<br>(0.0645) | 0.2475***<br>(0.0735) |
| 2014/15 | 0.2830***<br>(0.0477) | 0.3753***<br>(0.0567) | 0.2928***<br>(0.0645) | 0.3548***<br>(0.0736) |
| Constant | -5.5603***<br>(1.9742) | -1.6016<br>(2.2067) | 0.3790<br>(2.4500) | -1.0291<br>(2.5715) |
| Observations | 713 | 713 | 713 | 713 |

\* $p < 0.10$ , \*\* $p < 0.05$ , \*\*\* $p < 0.01$ . Results are reported using observed information matrix standard errors (OIM) and are given in parentheses. Exposure term, log of hospital beds has a coefficient of 1 in models (1), (2) and (3). Due to convergence issues, model (4) for delayed days due to major causes uses log of hospital bed-days as the exposure term.

**Table 9.11:** Robust Test for Exogeneity

|  | (1)<br>*Total* | (2)<br>*NHS* | (3)<br>*Major* | (4)<br>*Major (NHS)* |
| --- | --- | --- | --- | --- |
| Foundation Trust | 0.0631<br>(0.1454) | −0.0049<br>(0.1240) | 0.0693<br>(0.1386) | 0.0208<br>(0.1383) |
| % of Male Patients | 0.0062<br>(0.0184) | −0.0273<br>(0.0234) | −0.0209<br>(0.0278) | −0.0413<br>(0.0293) |
| % of Emergency Admissions | −0.0037<br>(0.0044) | −0.0058<br>(0.0072) | −0.0019<br>(0.0070) | −0.0040<br>(0.0098) |
| % of Patients Aged 60–74 | 0.0138<br>(0.0244) | 0.0358<br>(0.0328) | 0.0503<br>(0.0391) | 0.0594<br>(0.0422) |
| % of Patients Aged 75+ | −0.0016<br>(0.0098) | −0.0066<br>(0.0133) | −0.0020<br>(0.0151) | −0.0046<br>(0.0177) |
| Hospital Beds 200–399 | −0.8046***<br>(0.1958) | −1.3058***<br>(0.1204) | −0.2676*<br>(0.1550) | −1.1721***<br>(0.1462) |
| Hospital Beds 400–599 | −0.6481***<br>(0.2323) | −1.1578***<br>(0.2285) | 0.0826<br>(0.3138) | −0.9171***<br>(0.3096) |
| Hospital Beds 600–799 | −0.8717***<br>(0.2496) | −1.3225***<br>(0.2616) | −0.2192<br>(0.3441) | −1.1578***<br>(0.3375) |
| Hospital Beds 800–999 | −0.9891***<br>(0.2595) | −1.5113***<br>(0.2747) | −0.4290<br>(0.3588) | −1.3731***<br>(0.3506) |
| Hospital Beds 1000–1499 | −0.8485***<br>(0.2899) | −1.3713***<br>(0.3191) | −0.1967<br>(0.3980) | −1.1879***<br>(0.4059) |
| Hospital Beds 1500+ | −0.6226**<br>(0.2869) | −1.3510***<br>(0.3147) | −0.2354<br>(0.3906) | −1.2818***<br>(0.3958) |
| % Readmissions (16–74) | −0.0295<br>(0.0272) | −0.0651*<br>(0.0334) | −0.0479<br>(0.0353) | −0.0685*<br>(0.0381) |
| % Readmissions (75+) | 0.0073<br>(0.0226) | 0.0257<br>(0.0287) | 0.0180<br>(0.0347) | 0.0232<br>(0.0357) |
| Special Staff | −0.0134<br>(0.0219) | −0.0120<br>(0.0261) | 0.0103<br>(0.0304) | 0.0062<br>(0.0302) |
| Management | −0.0034<br>(0.0065) | −0.0084<br>(0.0075) | −0.0072<br>(0.0096) | −0.0072<br>(0.0088) |
| Estimated FE Residuals | 0.0002<br>(0.0016) | 0.0007<br>(0.0020) | 0.0001<br>(0.0024) | −0.0003<br>(0.0024) |
| 2012/13 | 0.0790**<br>(0.0393) | 0.1575***<br>(0.0455) | 0.0928*<br>(0.0504) | 0.1300***<br>(0.0485) |
| 2013/14 | 0.1299**<br>(0.0519) | 0.2592***<br>(0.0661) | 0.1714**<br>(0.0718) | 0.2552***<br>(0.0743) |
| 2014/15 | 0.2819***<br>(0.0639) | 0.4050***<br>(0.0745) | 0.2723***<br>(0.0835) | 0.3371***<br>(0.0845) |
| Observations | 752 | 744 | 748 | 740 |

\* $p < 0.10$ , \*\* $p < 0.05$ , \*\*\* $p < 0.01$ . The models are estimated using Poisson fixed effects estimation and reported using robust standard errors and given in parentheses. The effects of time invariant variables cannot be estimated due to within-effects estimation.

## Footnotes

1 In the reminder of this paper, the terms delayed discharges and transfers of care will be used interchangeably

2 NHS Trusts are also know as hospitals. However, a single NHS trust can run one or multiple hospitals. To prevent any ambiguities, these two terms will be interchangeably used throughout the reminder of this paper

3 Overlooking any possible nuanced differences in their meanings, in this paper the terms well–being, happiness and satisfaction are used interchangeably

